# Modeling the asymptomatic prevalence of SARS-CoV-2 epidemic in Italy and the ISTAT survey

**DOI:** 10.1101/2021.01.27.21250597

**Authors:** Marco Claudio Traini, Carla Caponi, Riccardo Ferrari, Giuseppe Vittorio De Socio

## Abstract

**objectives:** August 3rd, 2020, the Italian National Institute of Statistics (ISTAT) presented preliminary results of seroprevalence survey on the percentage of individuals affected by Covid-19. The survey aims to define (within the entire population of Italy) the portion of individuals that developed an antibody response against SARS-CoV-2. For the first time one has an estimate of the asymptomatic infected population and the possibility to acknowledge its potential rôle in the infection spread in Italy, one of the most affected areas in Europe. The information obtained allow a particularly sensitive validation of epidemiological models which include the asymptomatic class.

**methods:** The present study is devoted to the construction of a model able to simulate, in a systematic way, the asymptomatic group whose relevance in the, SARS-CoV-2 epidemic, has been recently investigated and discussed. The investigation involves the description of the first epidemic outbreak in Italy as well as the predictive analysis of the ongoing second wave. In particular the possible correction to the data of the serological tests because of their sensitivity and specificity.

**results:** The model: taken as an example of the models presently used, satisfactory reproduces the data of the ISTAT survey showing a relevant predictive power and relegating in a secondary position models which do not include, in the simulation, the presence of asymptomatic groups. The corrections due to the serological test sensitivity (in particular those ones depending on the symptoms onset) make the comparison between data and models less accurate.

**conclusions:** The predictions of the model confirm a relevant presence of asymptomatic individuals also during the second pandemic wave in Italy. The ratio of reported to unreported cases is predicted to be roughly 1:4. A more detailed knowledge of the results of the survey could allow to correct, in a relevant way, the data by means of the experimental evidences on the antibodies sensibility. The model analyses of the vaccination strategies, confirms the relevance of a massive administration with the beginning of the year to arrive at the end of the infection within August 2021.

## I. INTRODUCTION

Traditionally, epidemiological models have grouped people into two, three or four groups (compartments), usually denoted by Susceptible (*S*), Exposed (*E*), Infected (*I*), and Recovered (*R*). Contact between a member of the infected group *I* and another person belonging to the susceptible group *S* leads to the latter person becoming infected with a certain probability. Depending on the model, the susceptible person either becomes infected straightaway (SIR model), or enters an intermediate stage called Exposed (*E*) (SEIR model). In the latter scenario, it is assumed that contact between persons belonging to the *E* and *S* groups does not lead to fresh infections, because members of the *E* group do not carry a sufficient viral load to infect others through contact.

However, one of the characteristic features of the coronavirus pandemic is that many of the persons who contract the disease are “asymptomatic” (or group *A*). In fact the spread of severe acute respiratory syndrome coronavirus 2 (SARS-CoV-2) throughout the world has been extremely rapid suggesting the hypotheses of a crucial role played by these infected persons who remain asymptomatic even if contagious (Buitrago-Garcia, 2020). A recent paper (Oran, 2020) summarizes the available evidences on asymptomatic SARS-CoV-2 infection concluding that “*asymptomatic persons seem to account for approximately 40*% *to 45*% *of the SARS-CoV-2 infections, and that they can transmit the virus to others for an extended period of time, perhaps longer than 14 days*”. Oran and Topol (Oran, 2020) conclude with the need “*that testing programs include those without symptoms*”. Other references estimate the fraction of asymptomatic patients to be more than 50% (Mizumoto, 2020) or as high as 75% (Day, 2020). For this reason, asymptomatic patients remain “hidden” and cannot be identified except through tests of large portions of the population. An example is given in the study by Lavezzo, E., E. Franchin, C. Ciavarella, *et al*.(Lavezzo, 2020) reporting results of a detailed survey in the small town of Vo’ (near Padua, Italy) where on February 21st a lockdown was imposed in the whole municipality as a first outbreak in Italy. After the lockdown they found a prevalence of 1.2% (95% CI:0.8-1.8%). Notably, 42.5% (95% CI:31.5-54.6%) of the confirmed SARS-CoV-2 infections detected across two surveys were asymptomatic (that is did not have symptoms at the time of swab testing and did not develop symptoms afterwards). After that a number of studies and projects have been developed to identify the rôle of asymptomatic in the pandemic infection in various countries (e.g. Buitrago-Garcia, 2020; Guerriero, 2020; Peterson, 2020; Pollán, 2020; Snoeck, 2020; Well, 2020; Yanes-Lane, 2020).

In August 2020 preliminary results of a seroprevalence survey have been presented for a set of 64660 persons in Italy (ISTAT, 2020). The ISTAT survey aimed to define (within the entire population) the portion of persons that developed an antibody response against SARS-CoV-2. The survey adopted a methodology allowing the evaluation of the seroprevalence in the population also estimating the fraction of asymptomatic (or subclinical) infections and the differences for age groups, sex, localization etc. The results are still preliminary because involve a restricted number of tests, in particular those ones whose results have been reported before July 27. The post-stratified techniques adopted ^1^ (Little, 1993) allow the production of statistical estimates coherent with epidemic data (both at the international and local level). For the first time one has an estimate of the asymptomatic infected population and its influence in the infection spread in Italy, one of the most affected area in Europe.

The results of the ISTAT survey, enlarged to the entire Italian region, are summarized in table I. The analysis concludes that 1482377 individuals developed IgG antibodies against SARS-CoV-2. A level of seroprevalence of 2.5% (95% CI: 2.3-2.6%) to be compare with 244708 officially reported cases.

**TABLE I.** Preliminary results of the seroprevalence analysis in Italy (ISTAT, 2020). The amount of IgG positive tests corresponds to an infected population of (almost) 1.5 million of individuals in Italy, when the official reported cases are 244708 till July 27th, 2020.

| Region | IgG positive<br>(95% CI) |  | absolute<br>value | reported<br>cases |
| --- | --- | --- | --- | --- |
|  | lower<br>limit | upper<br>limit |  |  |
| Italy | 2.3 | 2.6 | 1482377 | 244708 |

Asymptomatic patients (*A*) differ from exposed patients (*E*) in one important respect. Unlike in traditional epidemiological models, contact between a person in the *A* group and another in the *S* group does lead to the latter getting infected, with a certain probability. In addition, as in other models, contact between a person in the *I* group and another in the *S* group also leads to the latter getting infected, with a similar probability. The seminal paper by Robinson and Stilianakis (Robinson, 2013) formulates and analyzes a preliminary model that captures the asymptomatic phenomenon *A* and can be called SAIR, since the corresponding model including exposed *E* group is usually called SEIR. The two models differ in many aspects from a mathematical point of view; see Ansumali, *et al*.for a recent analysis and detailed discussion (Ansumali, 2020).

Moreover it is possible to develop more refined models of the pandemic by introducing additional categories such as Quarantined, Healed, Ailing, Recognized (or Detected), Threatened, etc. (e.g. Park 2020; Giordano, 2020; Brogi, 2020). By introducing more categories, one will get a more realistic model of the disease progression. On the other hand, the number of parameters to be estimated increases drastically. The ideal trade-off between these two conflicting considerations remains and has to be considered on a case by case basis (Jia, 2020a; Kinoshita, 2020; Yu, 2020).

Aim of the present paper is the study of asymptomatic compartment population during the SARS-CoV-2 epidemic event in Italy taking advantage of the ISTAT survey. The model used assumes *eight* compartments (groups) including symptomatic detected and undetected, quarantined, asymptomatic, threatened (hospitalized) and recovered, a model recently proposed to discuss the role of measures against the Italian outbreak (Traini, 2020).

## II. METHODS AND ANALYSIS

### A. A time dependent quarantined model with isolation

The model we use in order to describe the time evolution of the Italian outbreak is an epidemiological model originally proposed by Tang *et al*.in order to study the Wuhan event (Tang, 2020. J. Clin Med., Tang 2020, Infect. Dis. Model.). This model incorporates appropriate compartments relevant to intervention such as quarantine, isolation, and treatment. The population is stratified in Susceptible (*S*), exposed (*E*), asymptomatic infected individuals (*A*), infected with symptoms (*I*), hospitalized in a large sense (detected infected) (*H*), and recovered (*R*). Further stratification includes quarantined susceptible (*S*_*q*_), and isolated exposed (*E*_*q*_) compartments (see Fig. 1) which describe the tracing procedures.

**FIG. 1.**
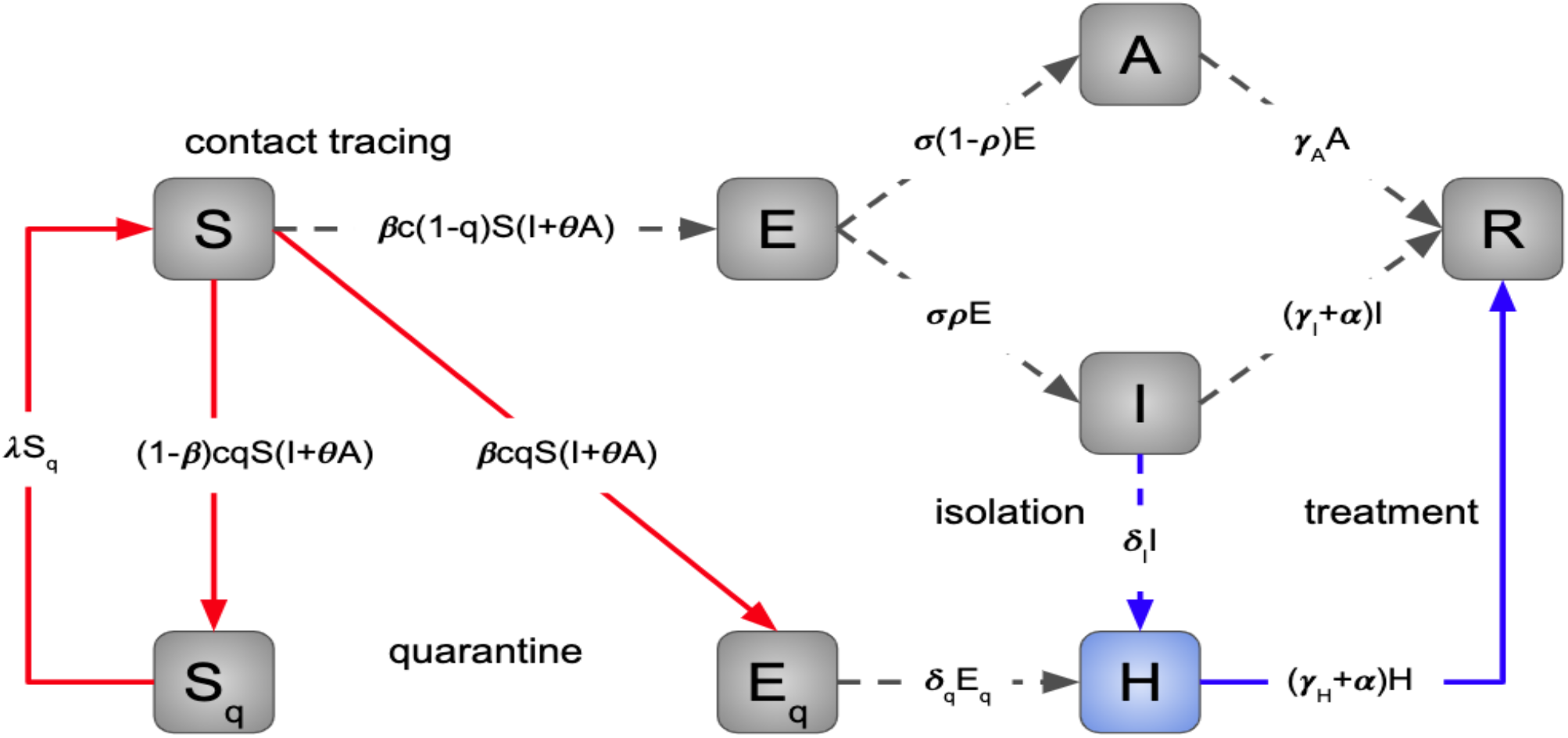
Diagram of the model simulating the novel Coronavirus (Sars-CoV-2) infection in Italy. The population is stratified in Susceptible (*S*), exposed (*E*), asymptomatic infected individuals (*A*), infected with symptoms (*I*), detected infected (hospitalized in a large sense) (*H*) and recovered (*R*), quarantined susceptible (*S*_*q*_), isolated exposed (*E*_*q*_) compartments. Interventions like intensive contact tracing followed by quarantine and isolation are indicated (Tang, 2020. Inf. Dis. Model.).

A portion of susceptible, *S*, get in contact with infected individuals with rate *c* (*I* + *θA*), where *c* is the contact rate, *I* the number of symptomatic infected individuals, *A* the asymptomatic infected individuals, and *θ* the contribution of asymptomatic infected to the infection spread. With contact tracing, a proportion, *q*, of susceptible, *S*, that get in contact with infected individuals is quarantined. Individuals receive the virus at rate *β*, which is the transmission probability, and become exposed. On the other side, the exposed individuals identified with contact tracing get quarantined at rate *q*. Therefore, we have three fluxes of individuals out of *S*: the quarantined with virus transmission going into *E*_*q*_, *βcqS*(*I* + *θA*); the quarantined without virus transmission going into *S*_*q*_, (1 − *β*)*cqS*(*I* + *θA*); the individuals with virus transmission but not identified and not quarantined going into *E, βc*(1 − *q*)*S*(*I* + *θA*). Quarantined individuals without virus transmission are released at rate *λ*, generating an inbound flux of individuals to *S* given by *λS*_*q*_. Exposed, infected, and quarantined individuals move to the hospitalized compartment at rate *δ*_*q*_*E*_*q*_. Exposed, infected, and not quarantined individuals become infectious at rate *σ*. Some of them develop symptoms with a probability of *ρ*. Then, there are two outbound fluxes of individuals for the compartment *E*: the infected with symptoms, *σρE*; the infected asymptomatic *σ*(1 − *ρ*)*E*. The infected with symptoms will eventually be detected and hospitalized with a rate of *δ*_*I*_, which reflects the sanitary system’s diagnostic capability. Finally, all the infected will recover with rate: *γ*_*A*_, for the asymptomatic, *γ*_*I*_ for the infected not hospitalized, *γ*_*H*_ for the infected hospitalized. The infected with symptoms, both hospitalized or not, have a mortality rate of *α*. For disease transmission, these individuals pass to the recovered compartment, as they are no more infectious. Summing the inbound and outbound fluxes at each compartment, we obtain the system of differential equations of the model:

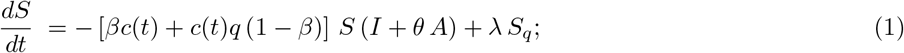

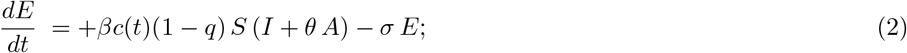

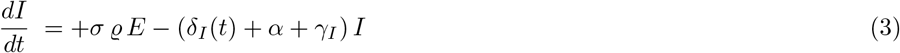

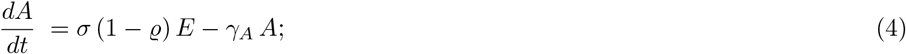

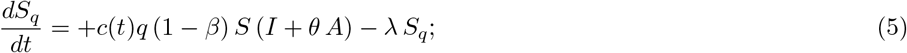

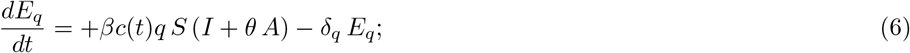

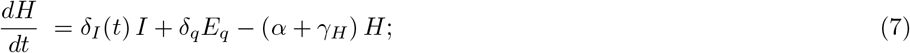

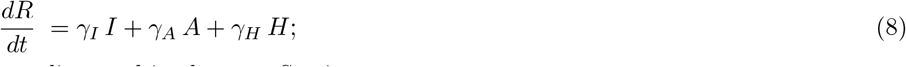

and the values of the parameters are discussed in the next Section.

### B. Fixing the model parameters: the outbreak

The method Markov Chain Monte Carlo (MCMC, (Brooks, 2011; Hogg, 2018)) is used to fit the model on the data of the outbreak in Italy in the period up to April 6th. The procedure is implemented through an adaptive Metropolis-Hasting (M-H) algorithm used for four concatenated runs with 100000 - 50000 - 25000 - 10000 iterations within the MCMC toolbox for Matlab. Table II summarizes the parameters. A peculiar aspect of the model is the time-dependence of the key parameters related to the contact rate *c* and the quarantined rate *q*. Following the control measures adopted in Italy, the flexibility of the model is such that one can adapt the values of some parameters to the concrete social situation. In particular the values *c*_0_, *c*_1_, *c*_2_ and *q*_0_, *q*_1_, *q*_2_ are fixed on the period of the outbreak February 24th - April 6th.

**TABLE II.** Parameters of the time-dependent model description.

| Param. | Definition [dimensions] | Estimated Value | St. Dev. | notes |
| --- | --- | --- | --- | --- |
| $c = c_0$ | Contact rate [$\text{day}^{-1}$] | 4.5248 | 0.2030 | from Feb. 24th to March 8th |
| $c = c_1$ | | $c_0/2$ | 0.2030 | from March 8th to March 29th |
| $c = c_2$ | | $1.4 \cdot c_1$ | 0.2030 | from March 29th to April 6th |
| $c = c_3$ | | $c_0$ | 0.2030 | from April 6th to Dec. 25th |
| $c = c_4$ | | $20 \cdot c_0$ | 0.2030 | after December 25th |
| $c = c(t)$ | see Section 2.3 | | | to discuss the lockdown exit of May 4th |
| $\beta$ | Probability of transmission per contact [u] | $1.5851 \cdot 10^{-8}$ | $3.360 \cdot 10^{-10}$ | |
| $q = q_0$ | Quarantined rate of exposed individuals [u] | $1.155 \cdot 10^{-7}$ | $7.720 \cdot 10^{-9}$ | from Feb. 24th to March 8th |
| $q = q_1$ | | $5.5 \cdot q_0$ | $7.720 \cdot 10^{-9}$ | from March 8th to March 29th |
| $q = q_2$ | | $2 \cdot q_1$ | $7.720 \cdot 10^{-9}$ | from March 29th to April 6th |
| $q = q_3$ | | $2 \cdot q_1$ | $7.720 \cdot 10^{-9}$ | from April 6th to Sept. 25th |
| $q = q_4$ | | $2 \cdot q_1$ | $7.720 \cdot 10^{-9}$ | from Sept. 25th |
| $\sigma$ | Transition rate of exposed individuals to the infected class [$\text{day}^{-1}$] | 1/7 | | |
| $\lambda$ | Rate at which the quarantined uninfected contacts were released into the wide community [$\text{day}^{-1}$] | 1/14 | | |
| $\varrho$ | Probability of having symptoms among infected individuals [u] | 0.2344 | 0.0060 | |
| $\delta_I = \delta_{I_0}$ | Transition rate of symptomatic infected individuals to the quarantined infected class [$\text{day}^{-1}$] | 0.1086 | 0.0010 | |
| $\delta_I = \delta_I(t)$ | see Section 2.3 | | | to discuss the lockdown exit of May 4th |
| $\delta_q$ | Transition rate of quarantined exposed individuals to the quarantined infected class [$\text{day}^{-1}$] | 0.1471 | 0.0010 | |
| $\gamma_I$ | Recovery rate of symptomatic infected individuals [$\text{day}^{-1}$] | 0.1704 | 0.0050 | |
| $\gamma_A$ | Recovery rate of asymptomatic infected individuals [$\text{day}^{-1}$] | 0.1177 | 0.0020 | |
| $\gamma_H$ | Recovery rate of quarantined infected individuals [$\text{day}^{-1}$] | 0.0515 | 0.0020 | |
| $\alpha$ | Disease-induced death rate [$\text{day}^{-1}$] | $1.2197 \cdot 10^{-5}$ | $3.0 \cdot 10^{-7}$ | |
| $\theta$ | Relative weight of asymptomatic infections [u] | 0.0840 | 0.0010 | |
| **Initial values** (February 24th) |  |  |  |  |
| $N$ | | $6 \cdot 10^7$ | | |
| $E(0)$ | | 21308 | 899 | |
| $I(0)$ | | 201 | 1.5 | |
| $A(0)$ | | 493 | 5.6 | |
| $S_q(0)$ | | 50000 | | |
| $E_q(0)$ | | 522 | 1.3 | |
| $H(0)$ | | 221 | | |
| $R(0)$ | | 8 | | |

In Fig. 2 some relevant predictions drawn from the model (together with some evident failures). The model is parametrized by means of the data on infected (reported cases) in the first time period (green data points in Fig. 2) till April 6th (day 42). It remains consistent with data till end of May (day 90), then it deviates from data. In fact the model was designed to study possible secondary effects after May 4th, when the lockdown in Italy has been gradually mitigated (see next Section II C). Its longterm behavior is discussed in detail (Traini, 2020). Here we want to emphasize an important result one can derive from the model predictions without formulating a new parametrization, namely the role of asymptomatic compartment as it emerges from the recent analysis by the Italian National Institute of Statistics (ISTAT, Istituto Nazionale di Statistica). As a matter of fact the model predicts also the global features of the distribution for the asymptomatic compartment illustrated in Fig. 2 and discussed in Sections III A and IV B.

**FIG. 2.**
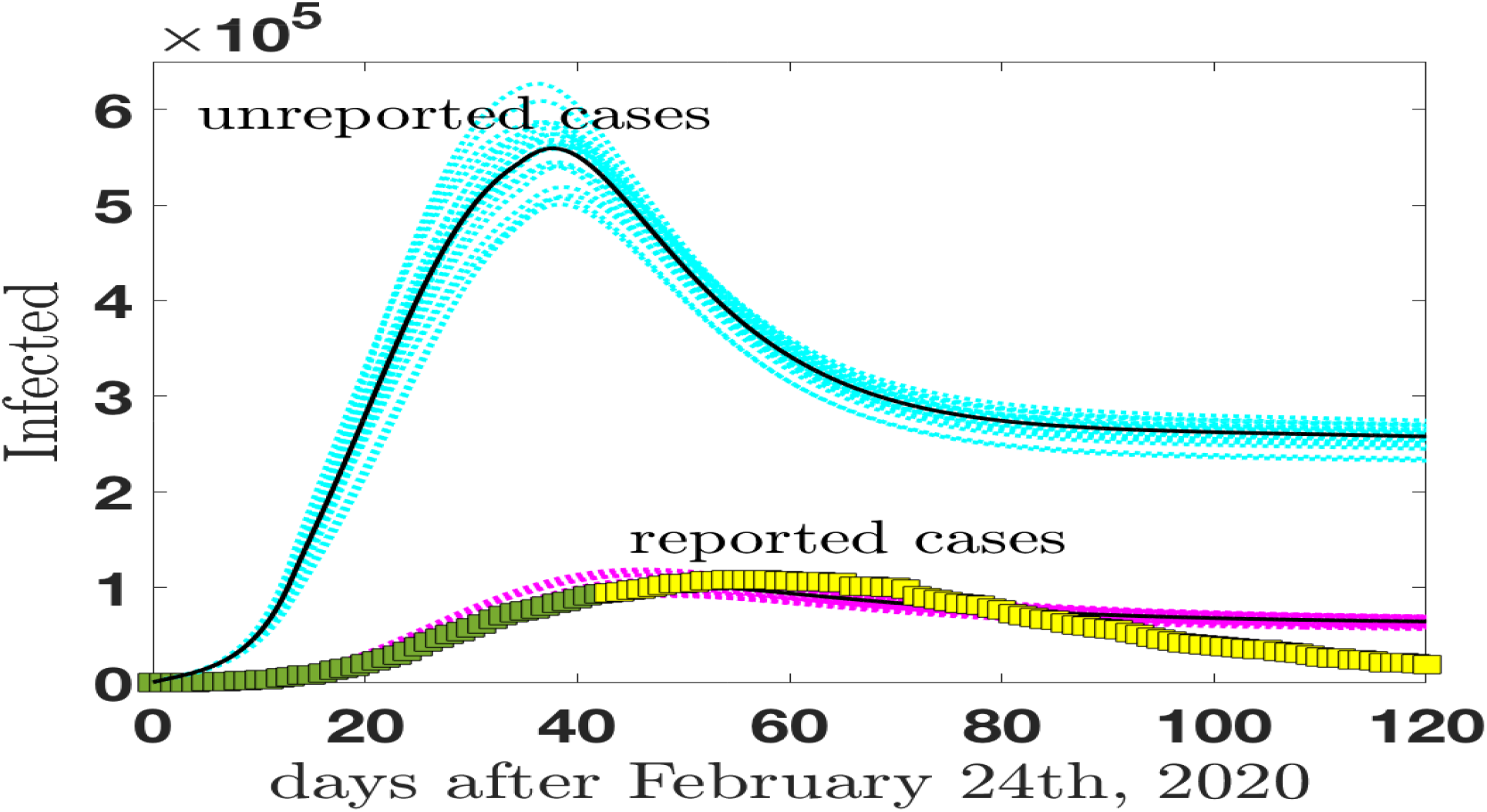
A MCMC analysis of the infected individuals (reported and unreported cases) in Italy as a function of time in the period February 24th - July 27th. The data for the reported-infected individuals are from (Ministero, 2020) and are compared with our model predictions (magenta curves) which include statistical uncertainties. The green data points refer to the data set used to fix the model parameters, while the yellow set spans the purely predicted period. The reported cases (magenta curves) are then compared with the predictions for the class of infected (asymptomatic and symptomatic) non reported cases (cyan curves) in the same period of time.

### C. predictive power: scenarios for secondary events

On March 8th, 2020, the Italian Government announced the implementation of restrictions for controlling the infection. Strong control measures (like convincing all the residents to stay home and avoid contacts as much as possible) have been adopted (lockdown).

From the model perspective, this can significantly contribute to decreasing the contact rate (*c*) among the persons (Remuzzi, 2020). Besides, the 2019-nCoV tests gradually shortened the time of diagnosis (i.e., the value of *δ*_*I*_ increased gradually). Considering these control strategies, we could tune the model on the concrete Italian conditions.

The equations of the model, shown in Eqs. (1)-(8), contain parameters explicitly dependent on time. In particular the contact parameter *c* and the transition rate of declared infected individuals *δ*_*I*_.

#### 1. time dependence

The initial Montecarlo analysis to fix the model parameters can be complemented with an accurate analysis of the effects of social measures or technological advances. An example is presented in Fig. 3 where the time behavior of the contact parameters and tracing rate is tuned following specific modeling related to concrete events changing the strength of the parameters involved. The time-dependence is parametrized as

**FIG. 3.**
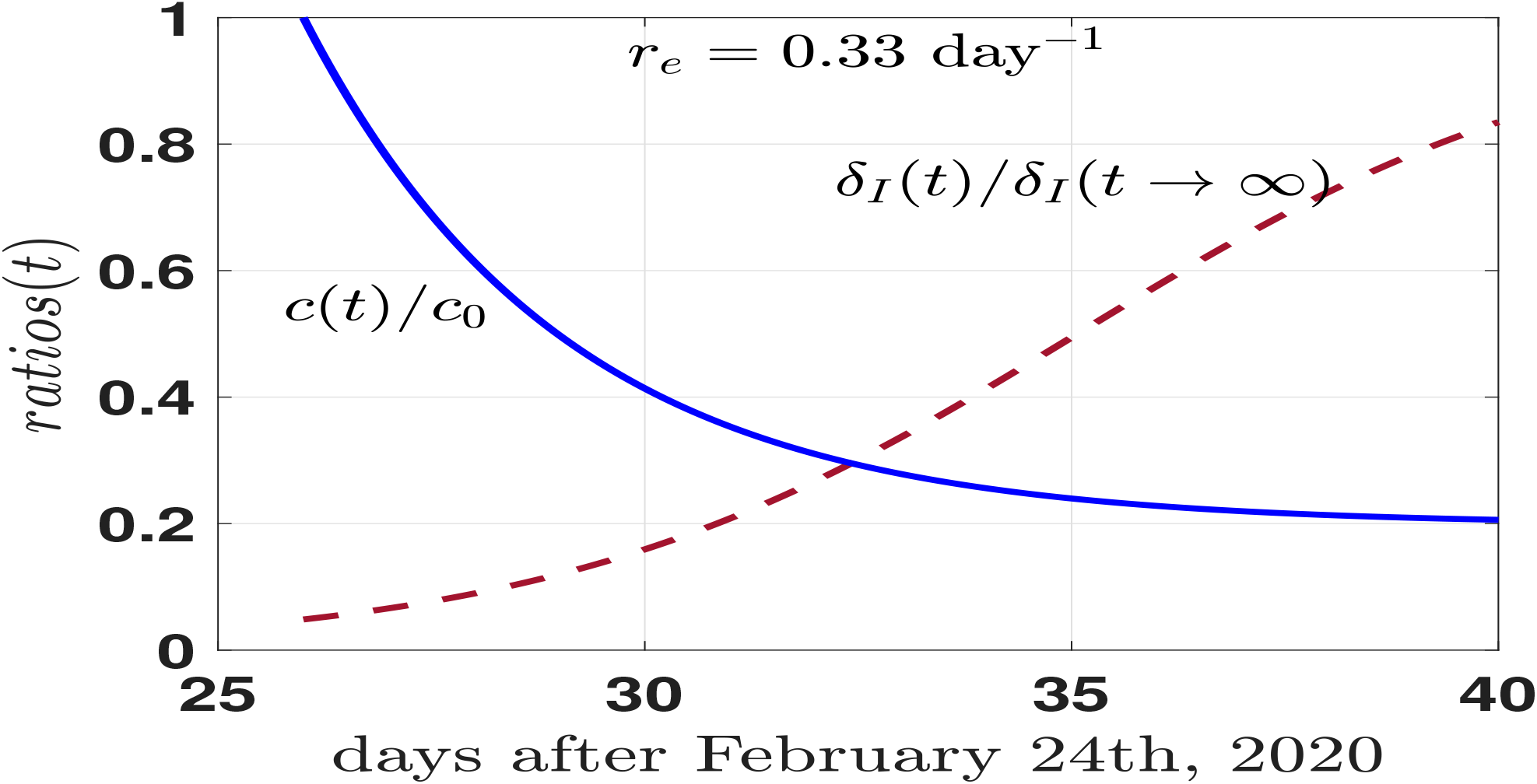
Modelling the variation in time of the contact rate per person *c*(*t*) (*c*(*t* = 0) = *c*_0_) and of the rate from infected to quarantined classes *δ*_*I*_ (*t*) (cfr. Eqs.(9),(10)).

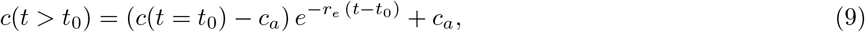

where *c*_*a*_*/c*_0_ = 0.2, *t* = 0 is fixed at February 24th and *t*_0_ selects the initial day of a rapid implementation of the isolation measures for the entire population. At the same time the parameter *δ*_*I*_, tuning the transition rate to quarantine of the infected individuals (*t*_*I*_ = 1*/δ*_*I*_), increases because of a decreasing of the testing time:

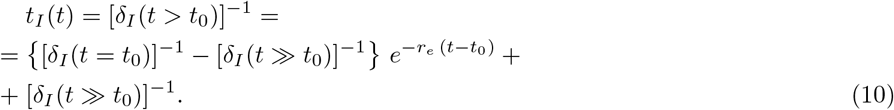

In Fig. 3 the behavior of the two parameters as a function of time. The rate of change *r*_*e*_ = 0.33 day^−1^ assumes the same values for the two gradually changing quantities. The parametrization (10), in particular, is chosen to simulate the behavior of the screening system to test infected (and asymptomatic) individuals.

#### 2. secondary events

Italy has been the first western country involved in the epidemic outbreak SARS-CoV-2. After 39 days (March 30th) the evolution was still in a first expanding phase. However, after almost 20 days of lockdown, the perspective of relaxing, at least partially, the social distancing measures appeared in many discussions at different levels and one can verify the predictive power of the model to simulate different scenarios of possible secondary effects. Let us assume that at a given day (the choice was May 1st, 2020), the containment measures are relaxed totally or partially. The time evolution of the contact rate could be described as in Fig. 4. The isolation value, as discussed in the previous Section, assumes the limiting value *c* ≈ 0.2 (isolation), and at day = 70th from February 24th, the isolation is interrupted and the “normalcy” activated. Two scenarios are introduced: i) a full return to the previous style of life (rather unlike, but it represents a reference point); ii) a more realistic scenario with half isolation.

**FIG. 4.**
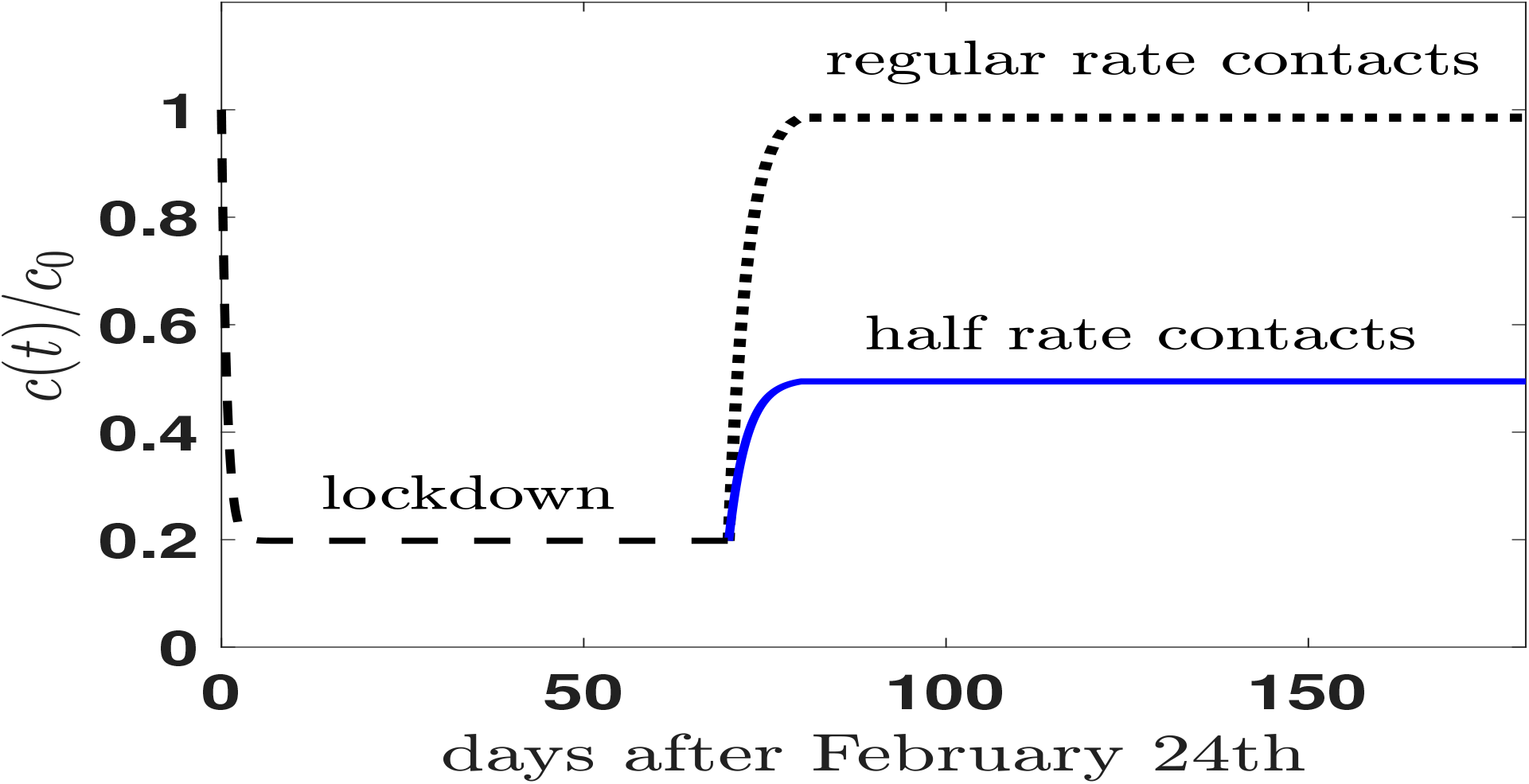
Time behavior of the contact rate simulating a possible secondary event in Italy at day=70, after February 24th, when the stringent measures of isolation are hypothetically relaxed. Two scenarios are introduced, the scenario of a rapid return to the old style of life (regular rate contacts), and a more realistic scenario where only half of the regular contacts are activated.

Assuming the parametrization of table II, one can try to reproduce the data on the reported cases. The good comparison between data and model can be valid for a short time, but we are not interested in reproducing the exact numbers, but the relative effects of a possible secondary event since the first outbreak did not exhaust its virulence. In addition, guided by the analysis of the effects of the technological developments in the screening phase done in the previous sections, one can try to see their effects in the secondary events and the results of such an investigation are shown in Fig. 5 where the comparison between the model predictions and the data is stringent. The data of the daily *new* (reported) cases, from February 24th on, show a large dispersion due to the influence of strategic decisions after the lockdown period (i.e. after May 4th). The behavior of the data follows rather consistently the model predictions of the reported cases of Eq. (7) once scenarios for the mitigation of the lockdown are introduced. In particular, the data seem to follow rather consistently a scenario (green lines) where the contact rate excludes a large part of social events (school, sports, large events) and it includes the introduction of tracing and quarantine. The fact that the longterm data (after May 4th) are well reproduced by the model means that distancing and tracing measures proposed in Italy had a positive impact captured by the model. More dramatic scenarios (as described, for instance, by the cyan curves) have been avoided. Particularly relevant the value of *t*_*I*_ = 1*/δ*_*I*_ reached during the transition time to normalcy. Lowering the diagnostic time to *t*_*I*_ ≈ 7 hours, or *t*_*I*_ ≈ 5 hours has the power to substantially mitigate the secondary events, in particular, if one keeps “half rate contacts” (as schematically designed in Fig. 4).

**FIG. 5.**
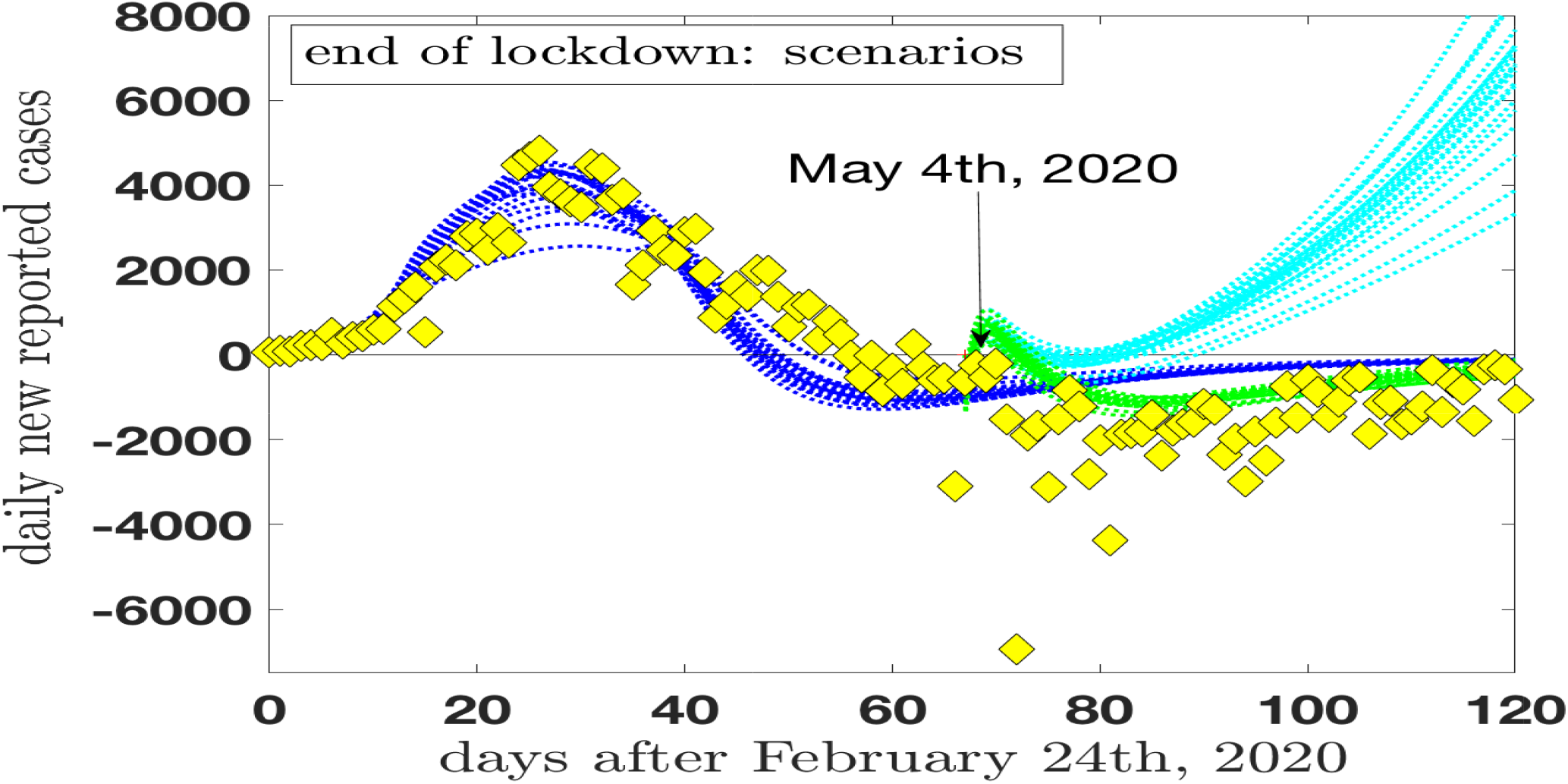
The model predictions for the daily new reported cases after February 24th, in Italy, are compared with the official data of ref. (Ministero,2020).. In addition to the fully locked scenario (blue lines), two other scenarios are included for a mitigation of the lockdown period in Italy starting from May 4th. Scenario A (cyan lines): a social contact rate somewhat similar to the regular rate (only a reduction of a factor of two is included) would result in a violent secondary outbreak. Scenario B (green lines): A stronger reduction of the contact rate which excludes a large part of social events (school, sports, large events…) with the further inclusion of tracing and quarantine.

## III. THE ASYMPTOMATIC PREVALENCE: RESULTS AND DISCUSSION

Epidemiological surveillance of COVID-19 cases captures only a portion of all infections because the clinical manifestation of infections with SARS-CoV-2 ranges from severe diseases, which can lead to death, to asymptomatic infection. The official sources of data in Italy (Ministero, 2020) do not provide information about the asymptomatic patients thus limiting their usefulness in view of calculating interesting epidemic parameters, not last the lethality rate. Attempts to exploit the existing available data in order to estimate the prevalence and the lethality of the virus in the total Italian population has been proposed by Bassi, Arbia and Falorsi (Bassi, 2020). They used a post-stratification (Little, 1993) of the official data in order to derive the weights necessary for re-weighting the sample results. The re-weighting procedure artificially modify the sample composition so as to obtain a distribution which is more similar to the population. They obtain a prevalence of 9%, a rather large number.

Conversely, a (population-based) seroepidemiological survey can quantify the portion of population which developed antibodies agains SARS-CoV-2 providing information on the exposed individuals and on the remaining susceptible subjects (assuming that antibodies are marker of total or partial immunity). A certain number of surveys of SARS-CoV-2 have been realized or planned (in addition to the already quoted references, one could mention Sood, 2020; Valenti, 2020; Stringhini, 2020; Bryan, 2020; Shakiba, 2020; Doi, 2020; Eriks 2020; Wu, 2020; N. Bobrovitz, 2020).

### A. Model results and ISTAT survey

The data, elaborated by ISTAT in their report, have been collected in a period of time (May 25th to July 15th) running over (almost) two months. They are summarized in the upper and lower limits and the absolute value of table I or (approximately) in the following ratio

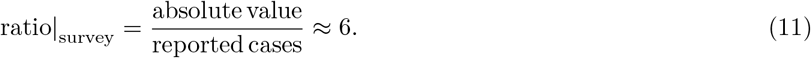

The ratio (11) is a clear sign of the large impact of the asymptomatic prevalence on the total infected population with positive IgG in Italy even if no time dependence is assumed for those data, an assumption that needs some more attention. In fact, the model description we are proposing can be of some help to answer questions on the time dependence of the data. In fact, one can presume a roughly constant behavior of the ratio (11) as a function of time under the following assumptions:

i. The antibodies have a lifetime significantly longer than the time needed for collecting the data.
ii. The social contact rate regime is rather stable during the collection of data. In this way the fraction of infected population is (on average) connected with the transmission probability of the infection which is assumed to be constant. In the present model *β* = (1.5851 *±* 0.0336) *·* 10^−8^ day^−1^, (cfr. table II).
iii. In a (large and representative) sample, the portion of individuals reached by the infection in the previous months is fixed (next section IV will be largely devoted to discuss this point).

The assumption i) sounds reasonable and the assumption ii) can be explicitly checked in our model by defining the following (time dependent) ratio

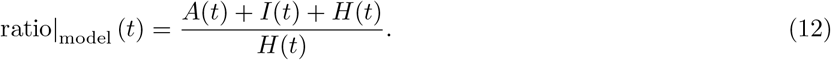

The numerator, in Eq. (12), counts, for each day, the total number of infected (reported and unreported with symptoms plus unreported asymptomatic infected individuals) and the denominator counts the number of reported cases. Both numerator and denominator are largely time dependent functions as already discussed in the previous sections. The model ratio (12) is shown in Fig. 6. Despite the rapid decrease of the ratio before May 25th, its value remains, after that date and till the end of July, approximately constant (ratio_model_(t) ≈ 5). In fact, the contact rate between February 24th and May 25th had large variation due to lockdown and social restriction measures adopted in Italy, while in the period May 25 - July 31st the social situation was rather stable. The model captures such variations validating the assumption ii) for that period.

**FIG. 6.**
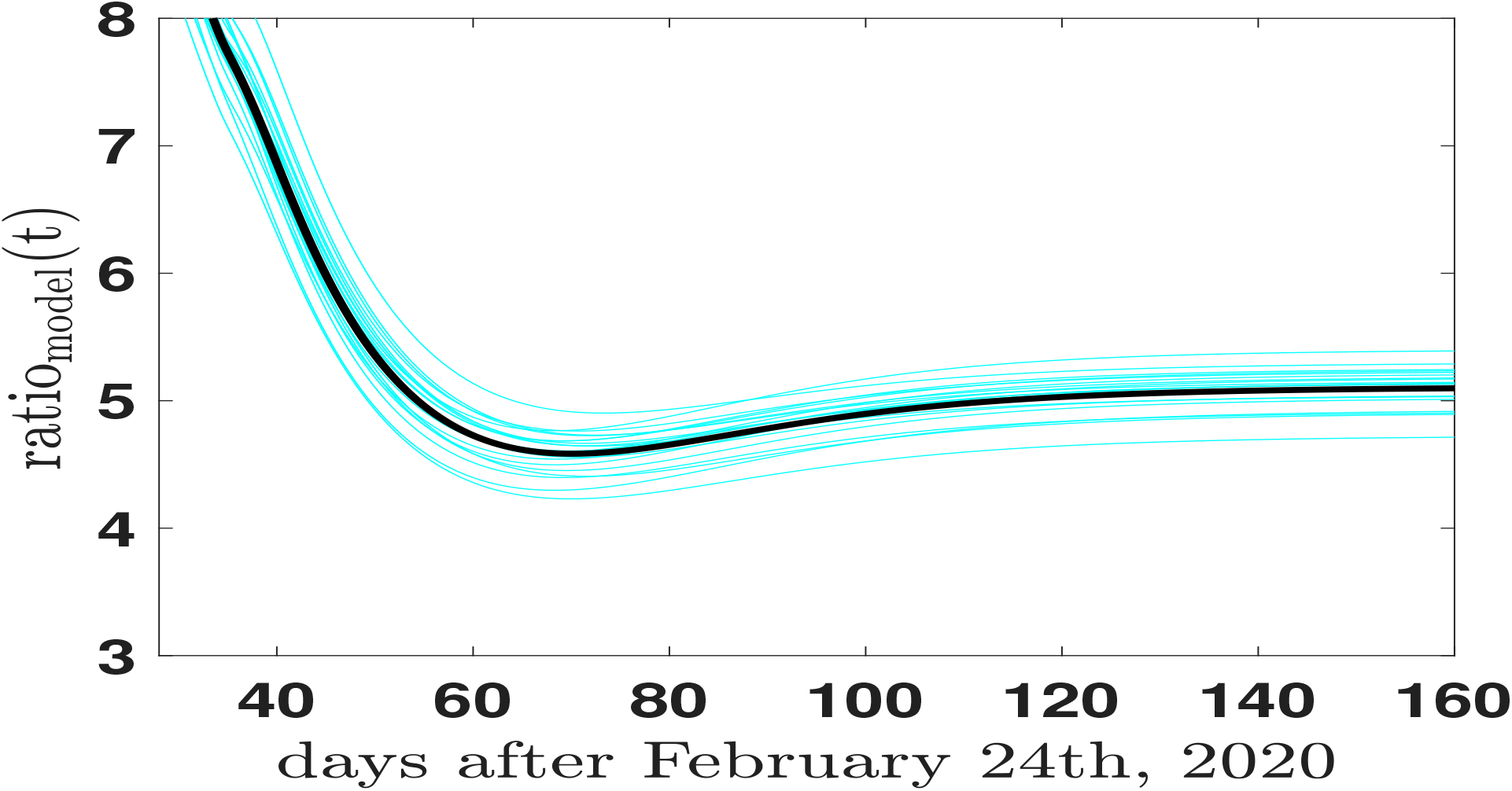
The model ratio of Eq.(12) as a function of time

However the ratio (12) cannot be compared directly with the results of the ISTAT survey. As a matter of fact the survey measured the number of individuals who developed antibodies without selecting the period of infection. The serological test cannot answer to time dependent questions only measuring the presence of antibodies which simply count the total amount of infections in the period preceding the test. In order to reproduce, as close as possible, the analysis from ISTAT, we can calculate the cumulative values of the numerator and denominator of Eq. (12) summing (up to a given day (*t*)) the components. The numerator of Eq. (11) (i.e. the absolute value) is given by the sum of the Asymptomatic population (*A*), the (unreported) Infected (*I*) and the official reported cases (*H*); the denominator the cumulative value of the official reported cases. One gets:

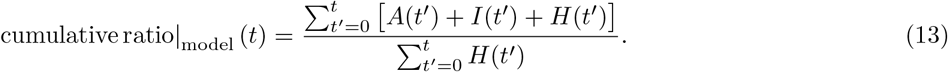

The cyan curves in Fig. 7 summarize the model predictions for the ratio (13). The black curve representing the mean value and the dotted lines the normal distributed variations.

**FIG. 7.**
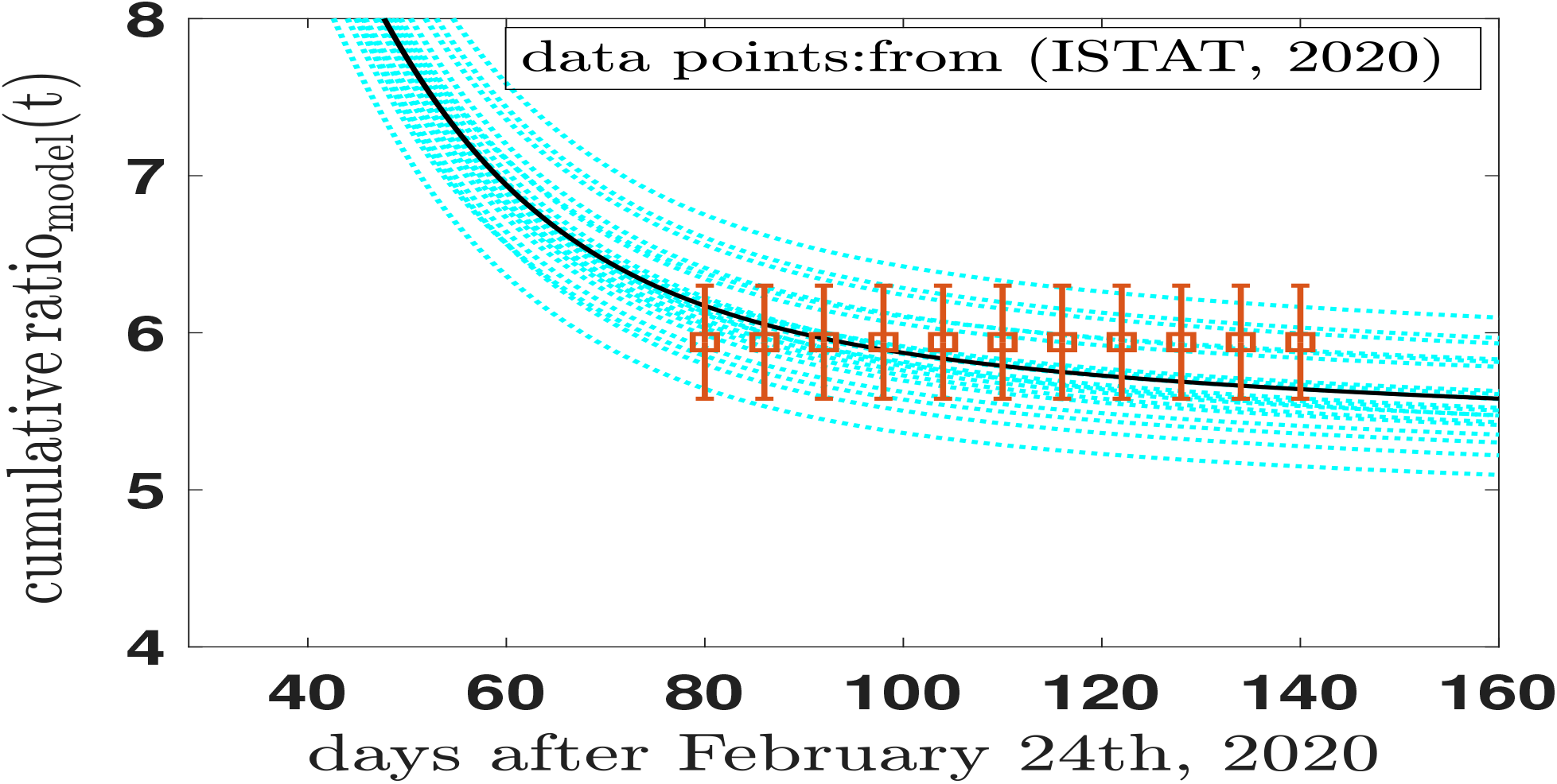
The fixed ratio of Eq.(14), elaborated from the results of the seroprevalence survey (data-points) is compared with the model ratio of Eq.(13) (cyan curves). See text for discussion.

The results of the ISTAT survey can be transformed in an analogous ratio in a simple way: the absolute value corresponds to an average value of the prevalence (2.3 + 2.6)*/*2 ≈ 2.5 (cfr. table I). Therefore the upper and lower limit of the infected population divided by the reported cases is summarized by the ratio

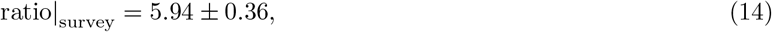

which now includes the uncertainties estimated in the ISTAT survey and updates the approximation (11). The data points in Fig. 7 give a graphical representation of the ratio (14) assumed, in a first step, to be time-independent on the basis of the assumption i) and ii) already discussed.

From this first analysis one can conclude that:

1. the global comparison in Fig. 7 between the estimated values and the evidences of ref. (ISTAT, 2020) is surprisingly successful, in particular when statistical uncertainties are included in the analysis of the survey and in the model predictions. For a long period of time during the lockdown months, speculations on the Asymptotic population gave strongly divergent estimates. Our results of Fig. 7 are obtained within a model fixed in April 6th, 2020 and published before the survey (Traini, 2020); data confirm that the epidemiological models can offer predictions rather stable and realistic also for the Asymptomatic compartment.
2. The model results for the cumulative ratio of Fig. 7 start with a value ≈ 8 at the beginning of April 2020, reaching an asymptotic *average* value ≈ 5.5 at the end of July 2020. For the whole period of the preliminary survey (May25 - July 15th) the ratio of Eq. (14), (data-points in Fig. 7), is reproduced by the model calculation of Eq. (13) within the statistical uncertainties.
3. The conclusions of the ISTAT survey, i.e.: “*the seroprevalence data at regional level, to be integrated with the epidemiological surveillance data, is particularly relevant to identify, on one side, the portion of individuals reached by the infection in the previous months, and for programming measures to prevent future possible second waves, on the other side”*^2^, can easily be applied to the relevance of modelling the behavior of the asymptomatic population, a key ingredient to manage the future of the pandemic event (e.g. (Gandhi, 2020)).

### B. Sensitivity and specificity of the serological tests

A first correction to the seroprevalence analysis presented in the previous section is due to the sensitivity and specificity of the serological tests adopted in the survey screenings (for a recent note on the false positive and false negative in diagnosis of Covid-19, see (Jia, 2020b)).

The report by ISTAT (ISTAT, 2020. page 9) indicates in *not less than 95%* the specificity of the tests and in *not less than 90%* their sensitivity. The consequent false negative and false positive classified individuals can be taken into account increasing the uncertainties of the ratio (14) of an amount of 10% and 5% in the two directions, assuming the maximum error in the data. One gets:

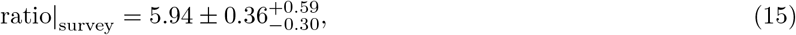

which replaces the value (14) to take into account the (asymmetric) corrections due to false responses. Fig. 7 is, consequently, replaced by Fig. 8 where the model results are compared with the more elaborated analysis. The asymmetric increase of the error-bars is clearly shown, however, at the same time, the conclusions of section III A remain basically valid.

**FIG. 8.**
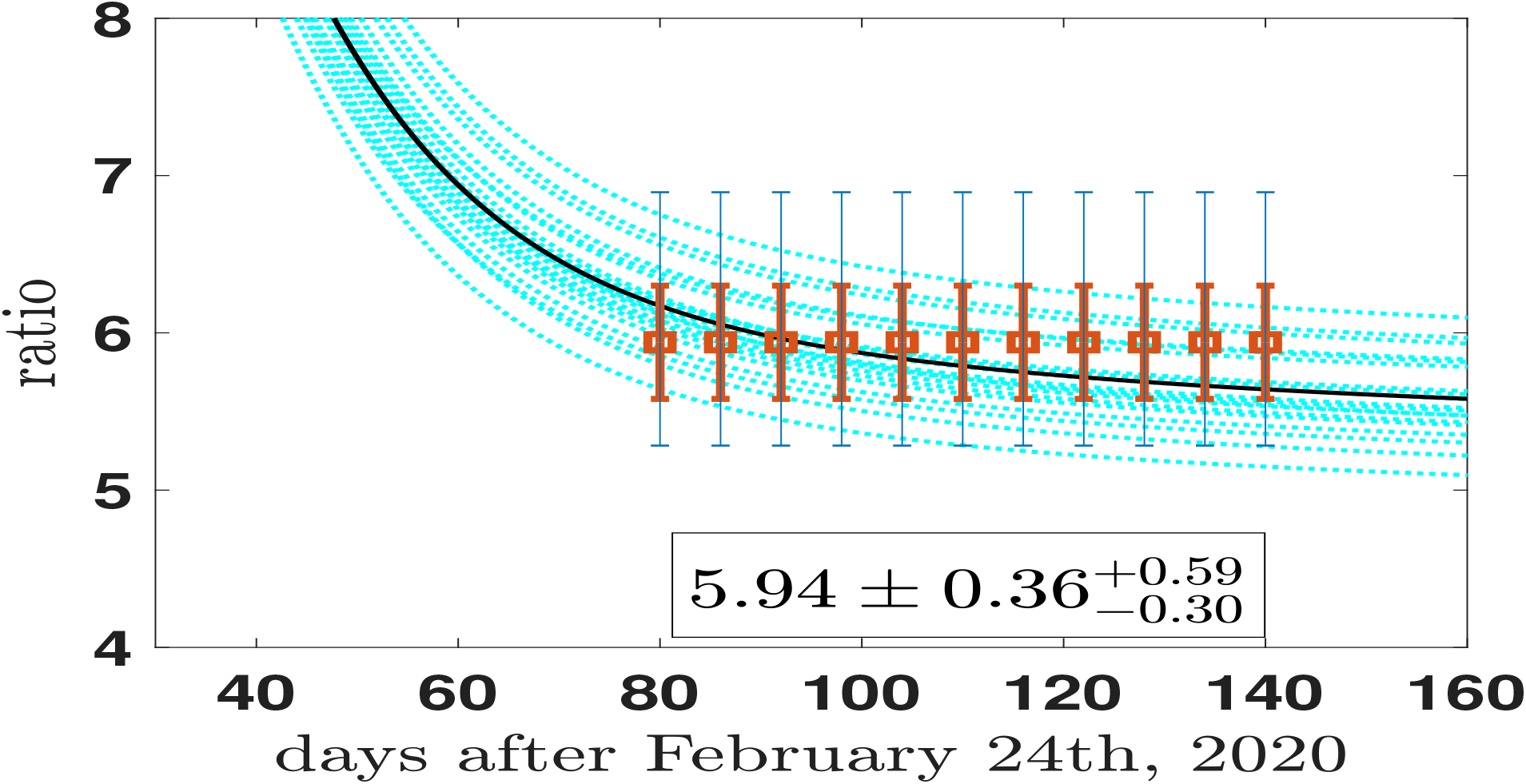
The ratios of Eqs.(14) and (15), are compared with the cumulative model ratio of Eq.(13) (cyan curves). The smallest error bars take into account the statistical variation suggested by the ISTAT report (ISTAT, 2020). The largest error bars include the modifications induced by the sensitivity and specificity of the IgG tests as indicated by the same ISTAT report. See text for discussion.

## IV. CRITICAL ASPECT AND LIMITS

The present section is devoted to the critical aspects of the results and methods used in the present investigation from two points of view: the data and the model. From the point of view of the data an important element is still not well fixed: the time dependence of the data during their collection, and it will be discussed in the next section IV A. From the point of view of the model study one cannot remain only with its analysis of the past and in section IV B a study of the present (and future) distribution of the reported cases and of the asymptomatic cases will be discussed. A stringent test for the model.

### A. Time dependence of antibody tests

A recent meta-analysis by Deeks *et al*.(Deeks, 2020) observed substantial heterogeneity in sensitivities of IgA, IgM and IgG antibodies, or combinations, for results aggregated across different time periods post-symptom onset. They based the main results of the review on the 38 studies that stratified results by time since symptom onset.^3^

In particular IgG/IgM all showed low sensitivity during the first week since onset of symptoms (all less than 30.1%), rising in the second week and reaching their highest values in the third week. The combination of IgG/IgM had a sensitivity of 30.1% (95% CI 21.4 to 40.7) for 1 to 7 days, 72.2% (95% CI 63.5 to 79.5) for 8 to 14 days, 91.4% (95% CI 87.0 to 94.4) for 15 to 21 days. Estimates of accuracy beyond three weeks are based on smaller sample sizes and fewer studies. For 21 to 35 days, pooled sensitivities for IgG/IgM were 96.0% (95% CI 90.6 to 98.3). There are insufficient studies to estimate sensitivity of tests beyond 35 days post-symptom onset. Summary specificities (provided in 35 studies) exceeded 98% for all target antibodies with confidence intervals no more than 2 percentage points wide. Assuming as a reference point the results of the meta-analysis by Deeks *et al*., one must correct further the uncertainties of Fig. 8. The questionnaire filled by the people involved in the screening for the ISTAT survey (ISTAT, Questionnaire 2020) included questions on the exact period of the symptom onset, a relevant information to correct the data and to analyze the effects of the time-dependent sensitivity of the tests. However the information are not at disposal at the moment and one is obliged to introduce more drastic assumptions and corrections. One remains with the simple assumption that sample has no privilege with respect the time dependence of the sensibility. As a consequence the single test has to be considered (in average) affected by the average value of the sensibility among 30.1% (days 1-7), 72.2% (days 8-14), 91.4% (days 15-21), 96% (days 22-35), and 90% for days ¿ 35 (and till the end of the screening period, i.e. the remaining 17 days). 90% is indeed the minimum value of the sensitivity proposed by ISTAT and discussed in section III B. One has:

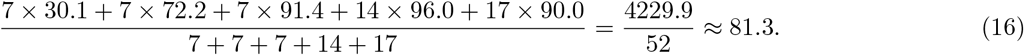

The sensitivity of the test decreases from *not less than 90%* to 81.3%, while the specificity remains *not less than 95%*.

Taking into account the new estimated sensitivity (16), the ratio (15) is replaced by

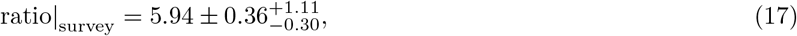

and the Fig. 8 by the Fig. 9. The asymmetric increase of the error-bars is again clearly shown, and the conclusions drawn in section III A become more weak.

**FIG. 9.**
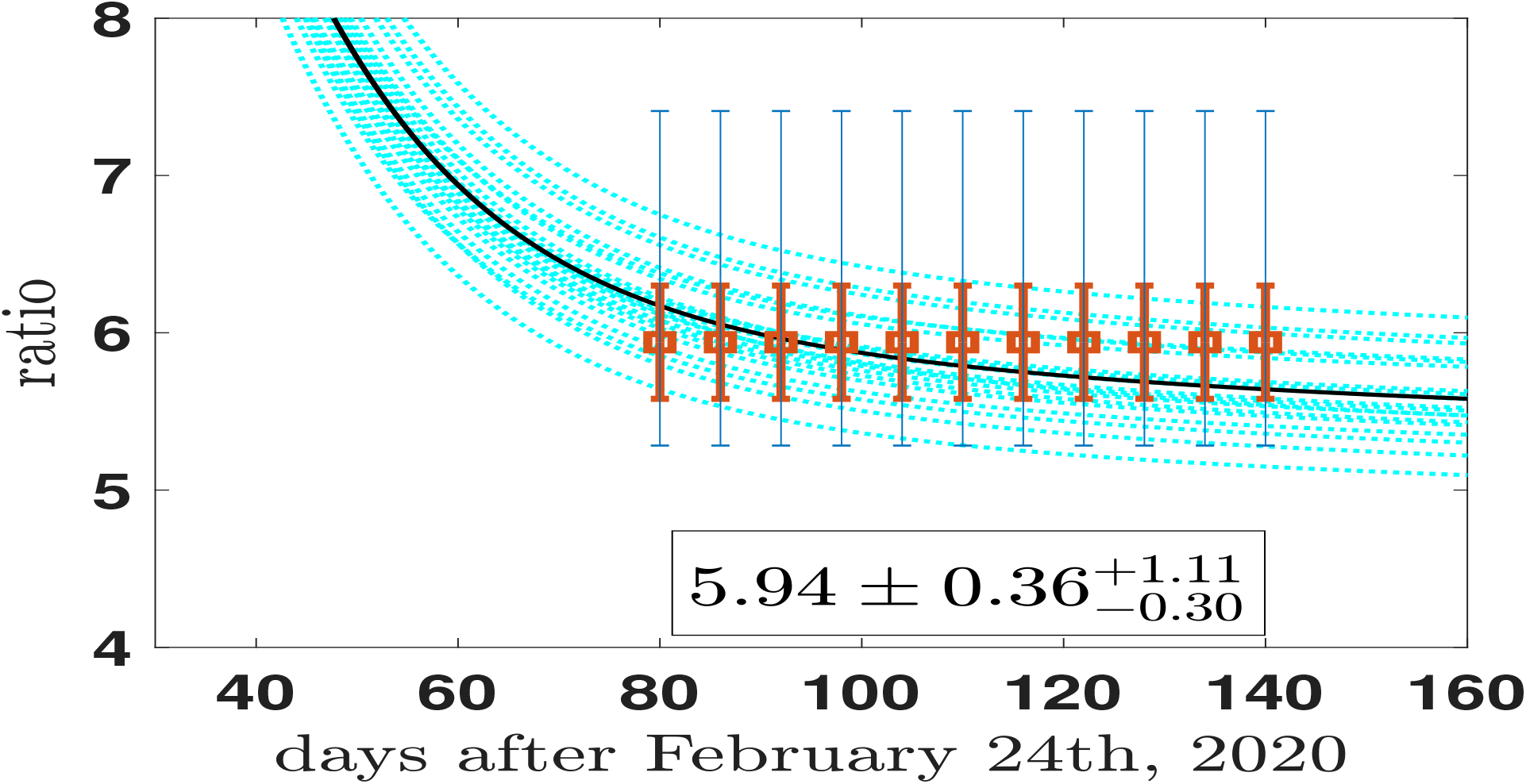
The ratios of Eqs.(14) and (17), are compared with the cumulative model ratio of Eq.(13) (cyan curves). The smallest error bars take into account the statistical variation suggested by the ISTAT report (ISTAT, 2020). The largest error bars include the modifications induced by the sensitivity and specificity of the IgG and their time-dependence as elaborated by Deeks *et al*.(Deeks, 2020). See text for discussion.

1. The comparison between the model estimates and the data corrected for the time dependence of the sensitivity of the tests as in Fig. 9 is less accurate and the data cannot be considered a stringent test for models. The estimate of the asymptomatic prevalence remains valid, but with a larger interval of confidence.
2. Model and corrected data are still consistent for the whole time period.
3. The validity of the model description remains a guide in the interpretation of the unknown asymptomatic distribution within a larger interval of values. A more careful analysis of the ISTAT data, in particular the reference to the mentioned questionnaire (ISTAT, Questionnaire 2020) could help in making again the comparison more stringent.

#### B. longer term study: last months of the year 2020

The present Section is devoted to the validation of the model (and predictions) with the most recent data on the second epidemic wave in Italy (after September 25th, 2020), in particular the predictions for the asymptomatic compartment. The parametrization is again based on values indicated in table II, no *ad hoc* modifications are introduced. The model is simply normalized to the value of the reported cases of September 25th (47718), as indicated in the official site of Ministero della Salute (Ministero, 2020). September 25th is assumed as the beginning of the second wave of infection. The new results are summarized in Fig. 10 (analogous to the Fig. 5 describing the previous period) and Fig. 11 (analogous to Fig. 2).

**FIG. 10.**
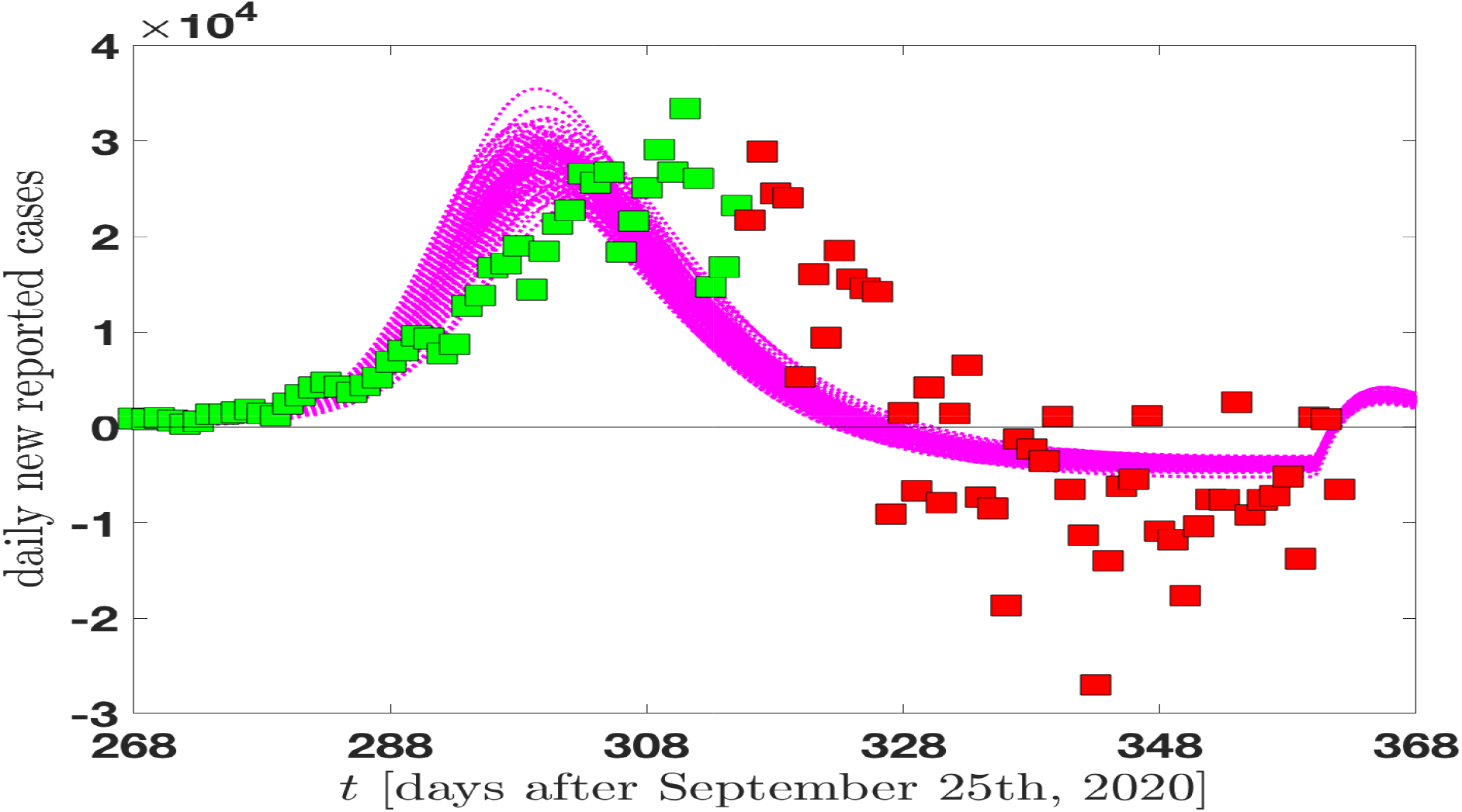
The model predictions for the daily new reported cases September 25th - December 31st, in Italy, are compared with the official data of Ministero della Salute (Ministero, 2020).

**FIG. 11.**
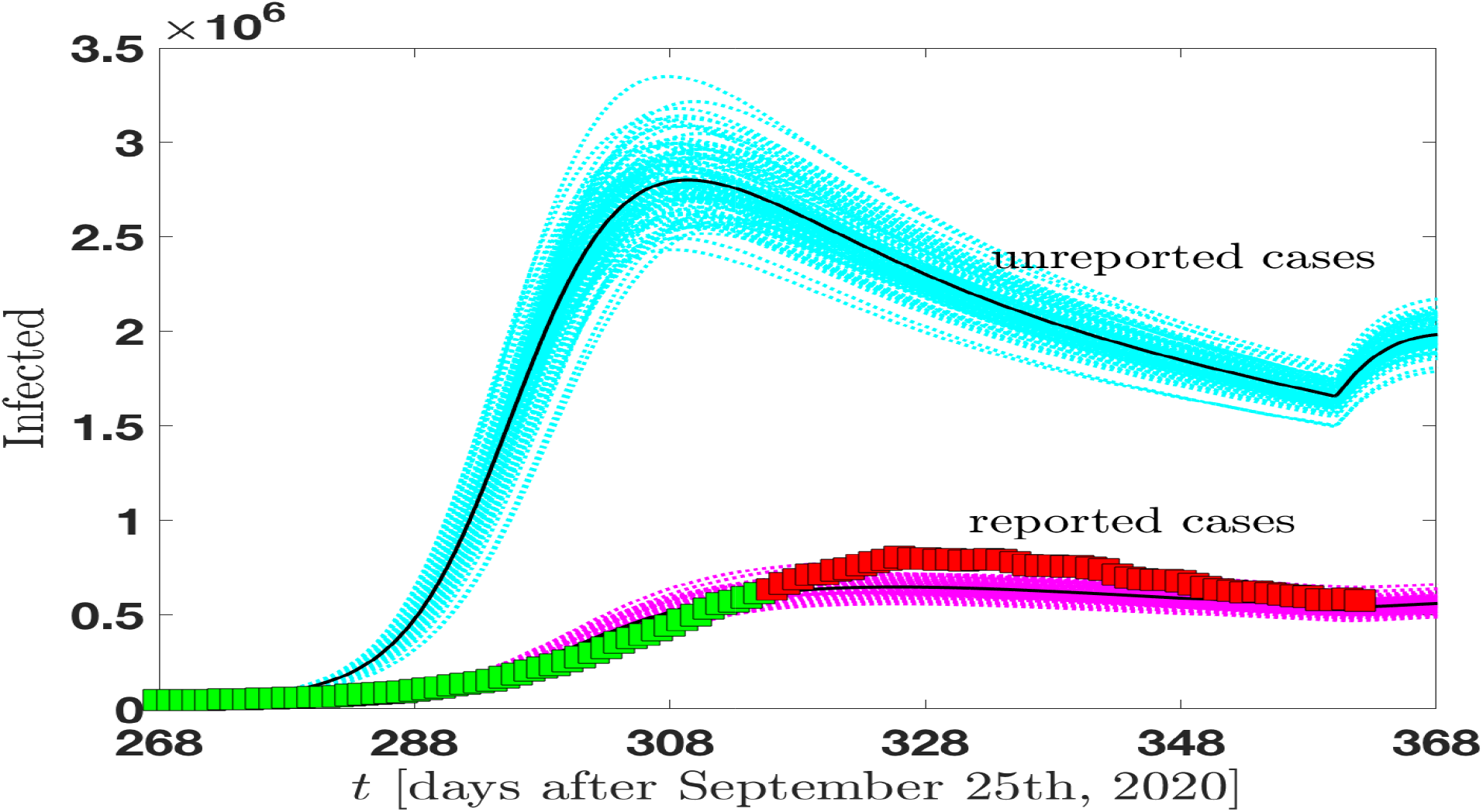
The MCMC analysis of the infected individuals (reported and unreported cases) in Italy as a function of time in the period September 25th - December 31st. The data for the reported-infected individuals are from (Ministero, 2020) and are compared with our model predictions (magenta curves) which include statistical uncertainties. The reported cases are compared with the predictions for the class of infected non reported cases (asymptomatic and symptomatic) in the same period of time (cyan curves).

The social measures adopted in the second period are rather different from the strict lockdown of the first events, in particular the measures assumed are locally differentiate and following the local virulence of the infection. The new approach of the governmental institutions assumes a strategically flexible response to the virus allowing for a possible “coexistence”. The response is therefore more rigid and without rapid variations. A behavior compatible with the results shown in Fig. 10 where the data for the daily variations exhibit a broader aspect in comparison with the model predictions. Despite such a differences, the model remain rather consistent with data of Fig. 10 which represent a stringent test since are related to the derivative of the distributions. In addition the data are submitted to large fluctuations due to data collection variation and local inhomogeneities. The data are presented in two different colors: green (till November 13th) and red afterwards (last update December 28th). The simple raison is that the model has been applied for the first time to the new data in November 13th. After November 13th the additional official data have been added to the Figure, day by day, and no modification or renormalization is introduced, i.e. the model predictions are fixed.

Much more stable are the results of Fig. 11 where data points from (Ministero, 2020) for the daily reported cases are shown as a function of time (days), from September 25th on… (last update December 28th). Once again the model predictions are compatible with data to a large extent. Such a consistency allows again a risky prediction for the unreported (asymptomatic and symptomatic) cases. The prevalence is rather large (approximately a factor of 4) quite similar to the prevalence emerging from the analogous Fig. 2 (factor of 5 roughly, see also Fig. 6). Obviously one needs detailed serological tests to have confirmation, however the stability of the virus is assumed by the parametrization of table II and seems to be consistent with data.

A last information emerges from Fig. 11: an indication of the possible effect due to the Christmas period. In Figs. 11 and 10 one can see the effect of a foreseeable relaxation of the distancing measures in the behavior of families and friends group. An average increase of the contact rate (as indicated in table II) produces a small wave emerging just after December 25th. A graphical indication of a possible increase of the number of infected.

## V. A LOOK AT VACCINATION

The section is devoted to a hot aspect of the nowadays ^4^ discussion on SARS-CoV-2: vaccination. Few days ago Great Britain started the vaccination campaign and on December 27th all the European countries will start their own campaign. The organization of an historical massive event is rather heavy and it will need the effort of many institutions.

### A. Modeling vaccination: scenarios

A preliminary aspect is the assessment of a possible vaccination scenario. We do not discuss specific strategies, but we want to establish general scenarios to illustrate main advantages and disadvantages within simple assumptions. Basically we will assume that the year 2021 will be devoted to vaccination in Italy and that the order of magnitude of the vaccinated persons per day is between 40000 and 80000 (to be selected with specific social and homogeneity criteria) (e.g. Makoul, 2020).

In the following two basic scenarios will be introduced, as illustrated in Fig. 12.

**FIG. 12.**
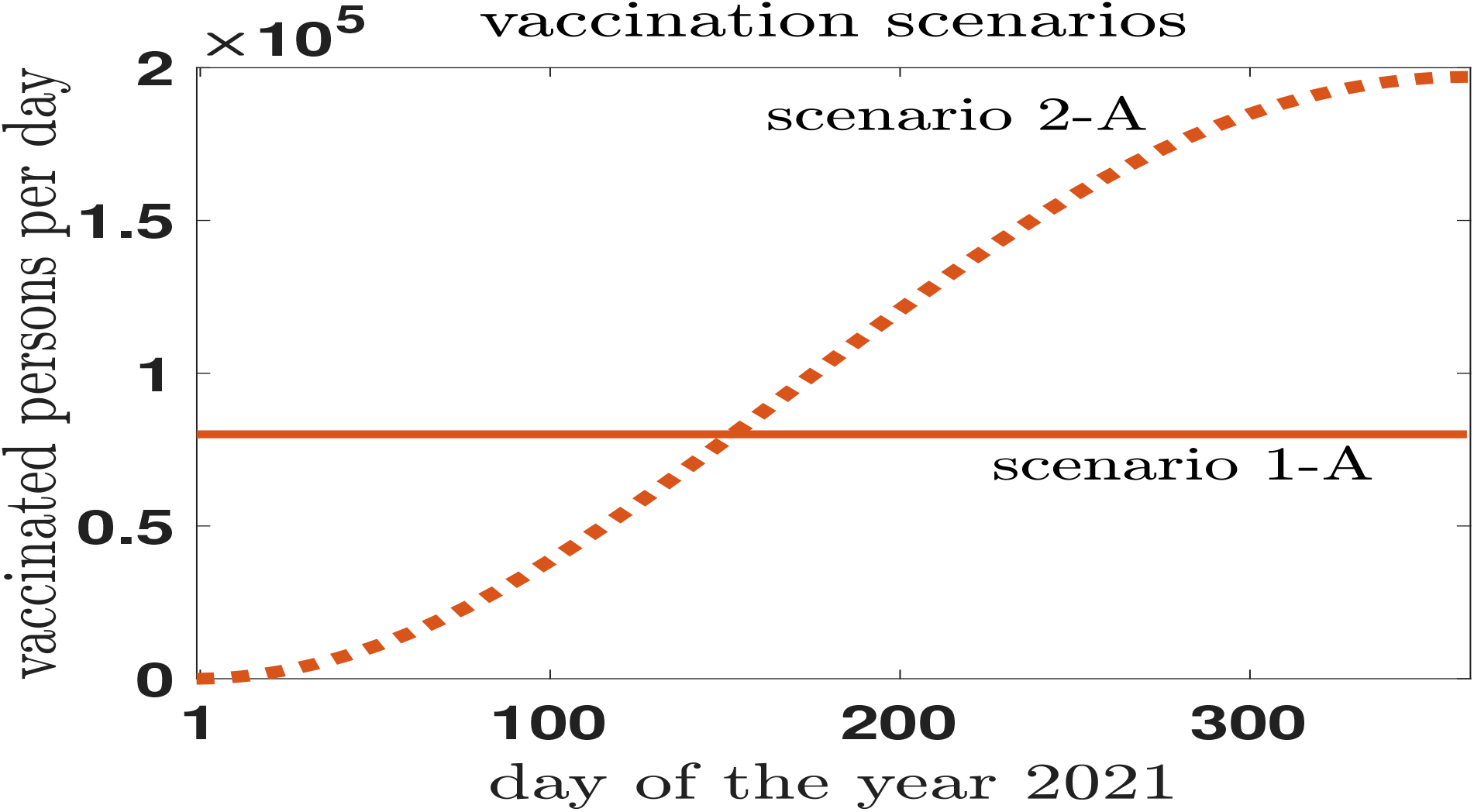
A constant number of person are vaccinated per day during the year 20121: 80000, full curve (scenario 1-A). Alternatively in scenario 2-A an increasing number of people are vacinated during 2021 allowing for a less strong starting effort. The total number of vaccinated people result (almost) 30 million for both scenarios, at the end of the year.

i. The first scenario assumes an homogenous distribution, during the 2021, of a fixed number of vaccinated person per day: 80000 (scenario 1-A), (full line in Fig. 12), and also 40000 (scenario 1-B) will be considered.. The effort to start with a large portion of population is high and a second scenario is investigated;
ii. The second scenario assumes that the amount of persons vaccinated during the first period is rather low and the process will accelerate during the year (dotted line in Fig. 12, scenario 2-A).
iii. Almost 30 million people are vaccinated at the end of the year in both scenarios.

### B. year 2021: a perspective

The section is devoted to a general perspective of the year 2021 offering a macroscopic view of the pandemic event in Italy and the advantages of the vaccination campaign. In Fig. 13 one can have an approximated idea of the time-evolution of the SARS-CoV-2 infection in Italy during the year ^5^ in the case of *no vaccination*, an assumption which is ruled out, but it can offer a reference point to appreciate the advantages of the vaccines. Such a scenario is described by the full black line of the upper panel (Fig. 13). One can immediately see that the number of the daily reported cases (today, December 28st, 2020, they are at the level of 575221) is decreasing during the year since a larger and larger number of “susceptible” individuals get immunized (or dead) after the infected period. The year 2021 will not be sufficient to obtain a vanishing number of infected: in summer they will be around 150000 and during the next Christmas period roughly 50000. A rough estimate which takes into account oscillations due to possible waves of infections ^6^ as it emerges from the wave behavior of the curve. The situation changes largely with the introduction of the vaccination at a rate of 80000 persons per day (brown curve, scenario 1-A). A reduction of roughly 100000 cases is already evident in April, while in summer (June) one reaches the 20000 cases instead of 150000, in August the virus will give a definitive CIAO. As a matter of fact one is gaining several months (from six to ten months) because of vaccination without counting the number of deaths …

**FIG. 13.**
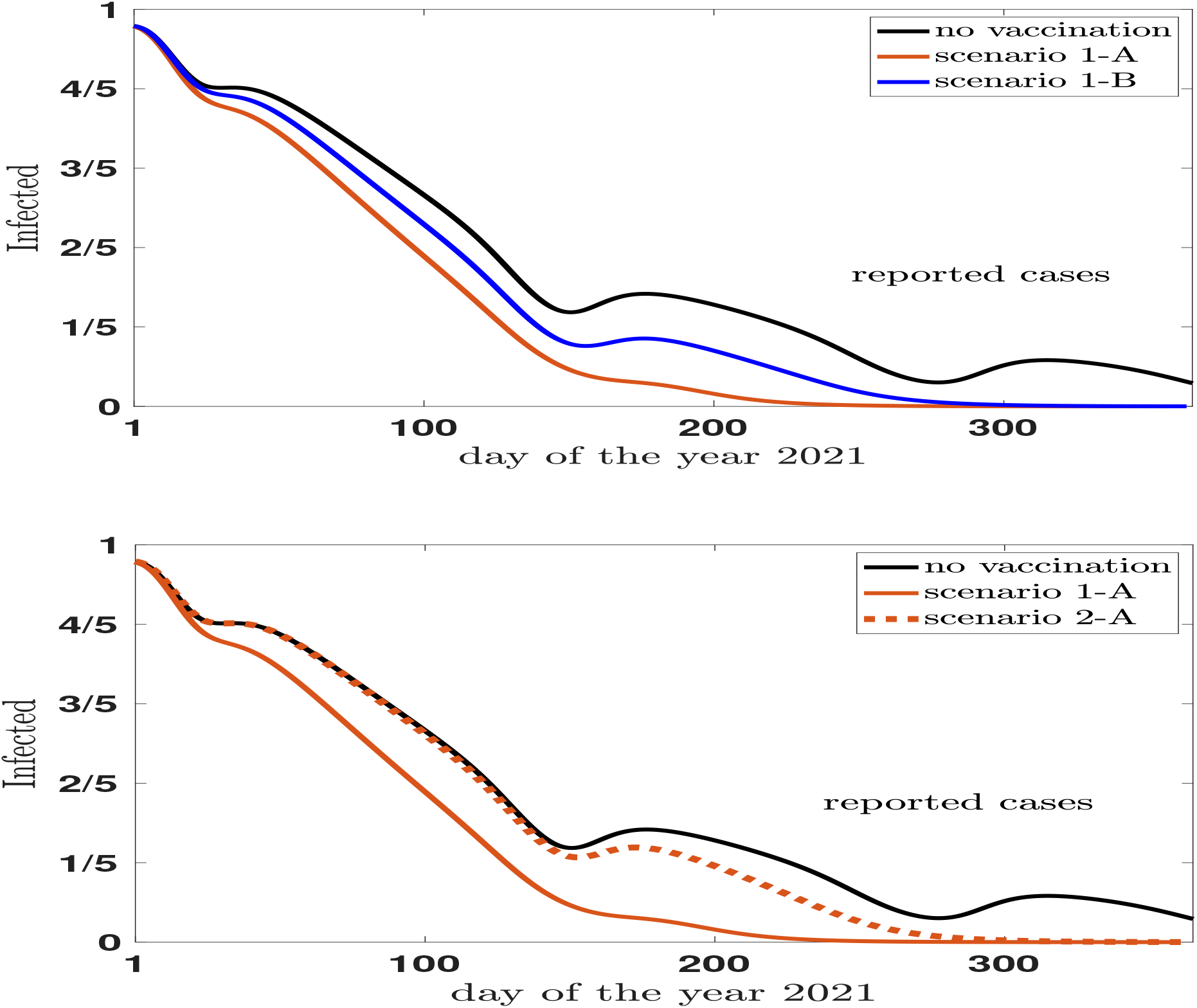
The fraction of the reported cases as a function of the day of the year 2021. **upper panel:** The scenario assuming no vaccination (continuous black line) is compared with the scenario 1-A which assumes a constant rate of vaccines of 80000 administrations each day (continuous brown line). The blu line (scenario 1-B) shows results for a administration of 40000 vaccines per day at constant rate. **lower pane:** The results of a constant rate of vaccine administration (80000 per day, continuous brown line) is compared with the results of no vaccination (black line) and the scenario 2-A where the rate is not constant and a low starting period is compensated by an accelerated second administration period (see Fig. 12).

The scenario 1-B (blu line, upper panel) assumes half vaccinated persons per day (40000) and the disadvantages are evident with respect the previous hypothesis of 80000. One has to wait end of October to eliminate the virus infection and in June one has still 80000 cases.

Also the scenario 2 has been implemented in the model and one can see the effects of a slow rate at the beginning of the year in the lower panel of the same Fig. 13. The results of the vaccination are almost invisible till the end of May. They appear rapidally in the second part of the year producing the final result at the end of October. The summer is still a hard period and scenario 2-A is rather similar to scenario 1-B.

Looking at Figs. 13 is not intuitive to accept that the reduction of the cases from 500000 to zero in one single year can produce the end of the infection in Italy with 60 million of habitants.

To partially restore intuition, one has to realize that, after the discussion of the asymptomatic cases of section III, a large part of the job is done by the unreported cases. In Fig. 14 the behavior of the total (unreported + reported cases) during the year as indicated by the model. They complement Fig. 13 showing that the decreasing behavior of the reported cases has a specular behavior for the asymptomatic. The vaccine helps in a strong way the reduction of both.

**FIG. 14.**
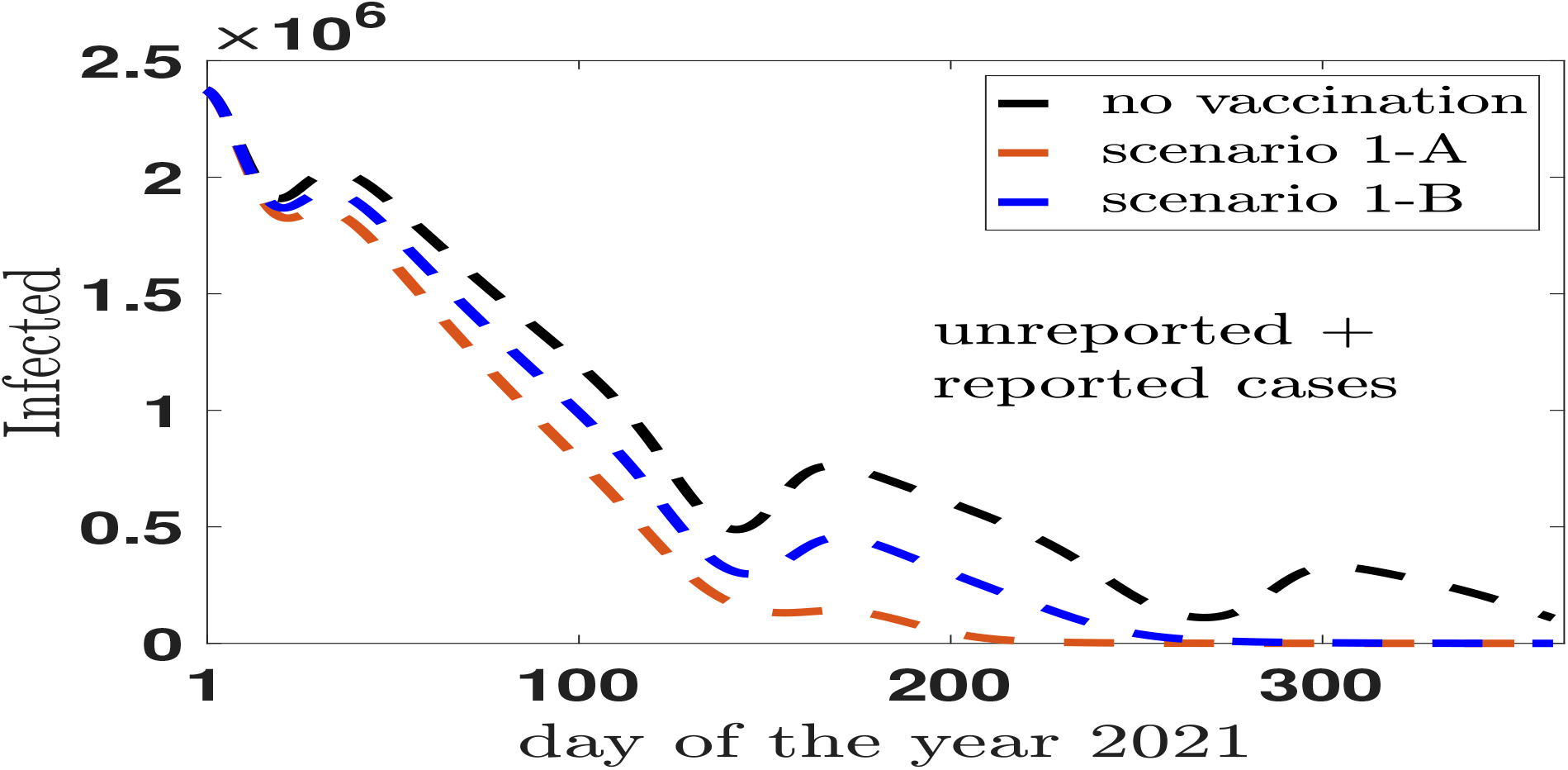
The reported+unreported cases as a function of the day of the year 2021. The scenario assuming no vaccination (dashed black line) is compared with the scenario 1-A which assumes a constant rate of vaccines of 80000 administrations each day (dashed brown line). The blu (dashed) line (scenario 1-B) shows results for a administration of 40000 vaccines per day at constant rate.

## VI. CONCLUDING REMARKS

The traditional compartmental classes of Susceptible, Exposed, Infected and Recovered which characterize a large fraction of epidemical models, are not sufficient to simulate the coronavirus (SARS-CoV-2) pandemic infection. Many of the people who contract the diseases are Asymptomatic, they are infected and contagious and are often invoked as one of the causes for the rapid spread of the infection. It is hard to estimate the amount of asymptomatic, estimates ranges from 40% to 75% (Buitrago-Garcia, 2020; Oran, 2020; Day, 2020; Muzimoto, 2020) of the total infected population. In a small scale the particular conditions of the Vo’ village (near Padua, Italy) allowed for two detailed surveys after a localized lockdown: the analysis found a prevalence of 1.2% (95% CI:0.8-1.8%). Notably, 42.5% (95% CI:31.5-54.6%) of the confirmed SARS-CoV-2 infections were asymptomatic (that is did not have symptoms at the time of positive swab testing and did not develop symptoms afterwards, (Lavezzo, 2020)).

The Italian National Institute of Statistics (ISTAT) presented, in August, preliminary results of seroprevalence survey on the percentage of individuals reached by the SARS-CoV-2 infection in the previous months. The survey aims to estimate, in a methodological precise way, also the asymptomatic infected population and its rôle in the infection spread in Italy, one of the most affected areas in Europe.

The present study investigates a model description of the entire infected population in Italy considering a eightfold compartmental model which includes Infected, Asymptomatic and Quarantined population in addition to the more classic Susceptible, Recovered, Exposed. The model is illustrated in few examples and used to investigate in detail the asymptomatic prevalence including also sensibility and specificity of the antibody tests used for the surveys. The results validate the model description and encourage other studies to detail, in a more quantitative way, the role of time-dependence of the sensitivity of the test used for the antibody screening.

Also predictions for the period of the second wave in Italy are presented and discussed, including asymptomatic predictions. Despite the fact the epidemiological surveillance of the second wave of the epidemic event in Italy is characterized by a strong use of swabs, the model predictions for the amount of unreported cases is of the same order of magnitude of the percentage already seen in the first outbreak. Future antibody screening will verify the present prediction.

To complete modeling, section V has been devoted to the vaccination scenarios. The rôle played by the introduction of vaccination is clearly shown in reducing in a significative way the time to reach the end of the infection. The strategies of the administration can be simulated and they favor an intense administration from the beginning. Starting with a low rate of administration risk to loose a large part of the advantage.

## Data Availability

data are from official sources (Ministero della Salute, Italy)
the numerical results are available on request

## Authorship contribution statement

The paper is the product of the joint work of the four authors.

## Acknowledgements

MCT and RF thank Francesca Greselin of the Department of Statistics and Quantitative Methods, University of Milano-Bicocca, for her interest and help in the elaboration of the model.

## Footnotes

1 Due to intentional and/or unintentional processes the characteristics of a sample may not represent the characteristics of the population of interest. To mitigate this potential bias survey researchers post-stratify base probability of selection survey weights so that sample characteristics match population control totals.

2 our translation. “I dati di siero-prevalenza a livello regionale, da integrare con quelli di sorveglianza pidemiologica, sono particolarmente preziosi sia per conoscere la quota di popolazione che è stata infettata nei mesi precedenti, sia per la messa a punto di programmi sanitari al fine di prevenire future ondate dell’epidemia e orientare adeguatamente le politiche sanitarie” (in Italian). (ISTAT, 2020. page 1.)

3 The numbers of individuals contributing data within each study each week are small and are usually not based on tracking the same groups of patients over time.

4 December 21st., 2020

5 The qualitative intent of the present discussion is put in further evidence by the fact that no-Covid deaths and newborns are not included in the investigation. One is simply assuming that they compensate approximately during the 2021. In addition we will not discuss vaccine efficacy assuming *V E* = 100% for two reasons: i) we do not know the real efficacy of the vaccines; ii) it is rather simple to rescale results for an accepted efficacy of 90% (Moghadas, 2020).

6 The oscillating behavior is simulated by an oscillating contact rate *c*(*t*) = c_0_ʱ0 *c*_0_ · cos^2^ (Ω*t*), with 0 ≤ t ≤ 365 and Ω = 4*π*=365, cfr. table II.

## References

Ansumali S., S. Kaushal, A. Kumar, M.K. Prakash, M. Vidyasagar. 2020. Modelling a Pandemic with Asymptomatic Patients, Impact of Lockdown and Herd Immunity, With Applications to SARS-CoV-2. Annual Reviews in Control (in press). doi: 10.1016/j.arcontrol.2020.10.003

Bassi, F., G. Arbia, P.D. Falorsi. 2020. Observed and estimated prevalence of Covid-19 in Italy: How to estimate the total cases from medical swabs data. Science of the Total Environment (in press). doi:10.1016/j.scitotenv.2020.142799

Bobrovitz, N., Arora R.K., Yan T. et al.. 2020. Lessons from a rapid systematic review of early SARS-CoV-2 serosurveys. medRxiv. doi:10.1101/2020.05.10.20097451

Brogi, F., B. Guardabascio, G. Barcaroli, 2020. Covid-19 in Italy: Actual Infected Population, Testing Strategy and Imperfect Compliance. doi:10.13140/RG.2.2.18275.50729.

Brooks S., A. Gelman, G. Jones, X.-L. Meng. 2011. Handbook of Markov Chain Monte Carlo CRC press.

Bryan A., G. Pepper, M.H. Wener, et al. 2020 Performance characteristics of the Abbott Architect SARS-CoV-2 IgG assay and seroprevalence in Boise, Idaho. J. Clin. Microbiol. 58. e00941–20. doi:10.1128/JCM.00941-20

Buitrago-Garcia D., D. Egli-Gany, MJ. Counotte, S. Hossmann, H. Imeri, A.M. Ipekci, et al.2020. Occurrence and transmission potential of asymptomatic and presymptomatic SARS-CoV-2 infections: A living systematic review and meta-analysis. PLoS Med 17. e1003346. doi:10.1371/journal.pmed.1003346

Day M. 2020. Covid-19: Identifying and isolating asymptomatic people helped eliminate virus in italian village. The BMJ. 165. doi:10.1136/bmj.m1165

Deeks J.J., J. Dinnes, Y. Takwoingi, C. Davenport, et al. 2020. textitAntibody tests for identification of current and past infection with SARS?CoV?2. Cochrane Database of Systematic Reviews. 6. doi:10.1002/14651858.CD013652

Doi A., K. Iwata, H. Kuroda, et al.2020. Estimation of seroprevalence of novel coronavirus disease (COVID-19) using preserved serum at an outpatient setting in Kobe, Japan: a cross-sectional study. medRxiv. doi:10.1101/2020.04.26.20079822

Erikstrup C., C.E. Hother, O.B.V. Pedersen, et al.2020. Estimation of SARS-CoV-2 infection fatality rate by real-time antibody screening of blood donors. Clin. Infect. Dis. ciaa849. doi:10.1093/cid/ciaa849

Gandhi, M., D. S. Yokoe, D. V. Havir. 2020. Asymptomatic Transmission, the Achilles’ Heel of Current Strategies to Control Covid-19 N Engl J Med. 382. 2158. doi: 10.1056/NEJMe2009758.

Garcia-Basteiro A.L., G. Moncunill, M. Tortajada et al.2020. Seroprevalence of antibodies against SARS-CoV-2 among health care workers in a large Spanish reference hospital. Nat Commun 11. 3500. doi:10.1038/s41467-020-17318-x

Giordano, G., F. Blanchini, R. Bruno, et al. 2020. Modelling the COVID-19 epidemic and implementation of population-wide interventions in Italy Nature Medicine 26. 855. doi:10.1038/s41591-020-0883-7

Guerriero M, Z. Bisoffi, A. Poli, et al.2020. Prevalence of asymptomatic SARS-CoV- 2-positive individuals in the general population of northern Italy and evaluation of a diagnostic serological ELISA test: a cross-sectional study protocol. BMJ Open 10:e040036. doi:10.1136/bmjopen-2020-040036

Hogg D.W., D. Foreman-Mackey. 2018. Data Analysis Recipes: Using Markov Chain Monte Carlo. The Astrophys. Journal Supp., 236. doi: 10.3847/1538-4365/aab76e

ISTAT, Italian National Institute of Statistics 2020. Primi risultati dell’indagine di seroprevalenza sul SARS-CoV-2. https://www.istat.it/it/files//2020/08/ReportPrimiRisultatiIndagineSiero-1.pdf. (August 3rd. 2020).

ISTAT, Questionnaire 2020.

https://www.istat.it/it/files//2020/05/Questionario-indagine-sieroprevalenza.pdf

Jia X., J. Chen, L. Li, N. Jia, B. Jiangtulu, et al. 2020a. Modeling the Prevalence of Asymptomatic COVID-19 Infections in the Chinese Mainland. The innovation. 1. 1000026. doi:10.1016/j.xinn.2020.100026?

Jia X., L. Xiao, Y. Liu, 2020b. False negative RT-PCR and false positive antibody testsConcern and solutions in the diagnosis of COVID-19. Journal of Infection. doi:10.1016/j.jinf.2020.10.007

Kinoshita, R., A. Anzai, S.-M. Jung, N.M. Linton, T. et al. 2020. Containment, Contact Tracing and Asymptomatic Transmission of Novel Coronavirus Disease (COVID-19): A Modelling Study. J. Clin. Med. 9. 3125. doi:10.3390/jcm9103125

Lavezzo, E., E. Franchin, C. Ciavarella, et al. 2020. Suppression of a SARS-CoV-2 outbreak in the Italian municipality of Vo’. Nature. 584. 425. doi:10.1038/s41586-020-2488-1

Little R.J.A., 1993. Post-Stratification: A Modeler’s Perspective. Journal of the American Statistical Association, 88. 1001. doi:10.1080/01621459.1993.10476368

Makoul M., H.A. Houssein, H. Chemaitelly, S. Seedat, et al.2020. Epidemiological Impact of SARS-CoV-2 Vaccination: Mathematical Modeling Analyses. Vaccines. 8. 668. doi:10.3390/vaccines8040668

Ministero della Salute. 2020. http://www.salute.gov.it/portale/nuovocoronavirus/

Mizumoto K., K. Kagaya, A. Zarebski, G. Chowell. 2020. Estimating the asymptomatic proportion of coronavirus disease 2019 (covid-19) cases on board the diamond princess cruise ship, Yokohama, Japan, 2020. Euro Surveillance. 25 (10) pp. 1. doi:10.2807/1560-7917.ES.2020.25.10.2000180

Moghadas S.M., T. N. Vilches, K. Zhang, et al. 2020. The impact of vaccination on COVID-19 outbreaks in the United States. medRxiv. doi:10.1101/2020.11.27.20240051.

Oran, D.P., E.J. Topol. 2020. Prevalence of Asymptomatic SARS-CoV-2 Infection. Ann. Intern. Med. June 3: M20-3012. doi:10.7326/M20-3012

Park, S.W., D. M. Cornforth, J. Dushoff, J. S. Weitz. 2020. The time scale of asymptomatic transmission affects estimates of epidemic potential in the COVID-19 outbreak, Epidemics 31. 100392. doi:10.1016/j.epidem.2020.100392

Peterson I., A. Phillips 2020. Three Quarters of People with SARS-CoV-2 Infection are Asymptomatic: Analysis of English Household Survey Data Clin. Epidemiology. 12. 1039. doi:10.2147/CLEP.S276825

Pollán, M., Párez-Gómez, B., Pastor-Barriuso, R., Oteo, J., et al.2020. Prevalence of SARS-CoV-2 in Spain (ENE- COVID): a nationwide, population-based seroepidemiological study. Lancet. 396. 535. doi:10.1016/S0140-6736(20)31483-5.

Remuzzi, A., G. Remuzzi. 2020. COVID-19 and Italy: what next?. Lancet. 395. 1225. doi:10.1016/S0140-6736(20)30627-9

Robinson R., N.I. Stilianakis. 2013. A model for the emergence of drug resistance in the presence of asymptomatic infections. Mathematical Biosciences. 243. 163. doi:10.1016/j.mbs.2013.03.003

Salje H., C.T. Kiem, N. Lefrancq, et al.2020. Estimating the burden of SARS-CoV-2 in France. Science. 369. 208. doi:10.1126/science.abc3517

Shakiba M., S.S.H. Nazari, et al.2020. Seroprevalence of COVID-19 virus infection in Guilan province, Iran. medRxiv. doi:10.1101/2020.04.26.20079244

Snoeck C.J., Vaillant, T. Abdelrahman, V. P. Satagopam, et al.2020. Prevalence of SARS-CoV-2 infection in the Luxembourgish population: the CON-VINCE study. medRXiv. doi:10.1101/2020.05.11.20092916

Sood N., Simon P Ebner P et al.2020. Seroprevalence of SARS-CoV-2-specific antibodies among adults in Los Angeles County, California. on April 1011, 2020. JAMA. 323. 2425. doi:10.1001/jama.2020.8279

Stringhini S., A. Wisniak, G. Piumatti et al.2020. Repeated seroprevalence of anti-SARS-CoV-2 IgG antibodies in a population-based sample from Geneva, Switzerland. The Lancet. 396. 313. doi:10.1016/S0140-6736(20)31304-0

Tang, B., X. Wang, Q. Li et al.2020. Estimation of the Transmission Risk of the 2019-nCoV and Its Implication for Public Health Interventions. J Clin Med. 9. 462. doi:10.3390/jcm9020462

Tang B., N.L. Bragazzi, Q. Li, S. Tang, Y. Xiao, J. Wu. 2020. An updated estimation of the risk of transmission of the novel coronavirus (2019-nCov). Infect Dis Model. 5. 248. doi:10.1016/j.idm.2020.02.001

Traini, M.C., C. Caponi, R. Ferrari, G. V. De Socio. 2020. A study of SARS-CoV-2 epidemiology in Italy: from early days to secondary effects after social distancing. Infectious Diseases 52. 866. doi: 10.1080/23744235.2020.1797157.

Valenti L., A. Bergna, S. Pelusi et al.2020. SARS-CoV-2 seroprevalence trends in healthy blood donors during the COVID-19 Milan outbreak. medRxiv. doi:10.1101/2020.05.11.20098442

Well P.M., J.K. Doores, S. Couvreur, R. M.a Nunez, et al.2020. Estimates of the rate of infection and asymptomatic COVID-19 disease in a population sample from SE England. Journal of Infection (in press). doi:10.1016/j.jinf.2020.10.011

Wu X., B. Fu, L. Chen, Y. Feng, 2020. Serological tests facilitate identification of asymptomatic SARS-CoV-2 infection in Wuhan, China. J. Med. Virol. 92. 1795. doi:10.1002/jmv.25904

Yanes-Lane M., N. Winters, F. Fregonese, M. Bastos, S. Perlman-Arrow, J.R. Campbell, et al.2020. Proportion of asymptomatic infection among COVID-19 positive persons and their transmission potential: A systematic review and meta-analysis. PLoS ONE 15. e0241536. doi:10.1371/journal.pone.0241536

Yu X., 2020. Modeling return of the epidemic: Impact of population structure, asymptomatic infection, case importation and personal contacts. Travel Medicine and Infectious Diseases 37. 101858 doi:10.1016/j.tmaid.2020.101858

